# Structural models of genome-wide covariance identify multiple common dimensions in autism

**DOI:** 10.1101/2022.10.21.22281213

**Authors:** Lucía de Hoyos, Maria T Barendse, Fenja Schlag, Marjolein MJ van Donkelaar, Ellen Verhoef, Chin Yang Shapland, Alexander Klassmann, Jan Buitelaar, Brad Verhulst, Simon E Fisher, Dheeraj Rai, Beate St Pourcain

**Affiliations:** Language and Genetics Department, Max Planck Institute for Psycholinguistics, Nijmegen, The Netherlands; Department of Social Dentistry and Behavioural Sciences, Academic Centre for Dentistry Amsterdam (ACTA), Amsterdam, The Netherlands; MRC Integrative Epidemiology Unit, University of Bristol, Bristol, United Kingdom; Population Health Sciences, University of Bristol, Bristol, United Kingdom; Institute for Genetics, University of Cologne, Cologne, Germany; Donders Institute for Brain, Cognition and Behaviour, Radboud University, Nijmegen, The Netherlands; Karakter Child and Adolescent Psychiatry University Centre, Nijmegen, The Netherlands; Department of Cognitive Neuroscience, Radboud University Medical Center, Nijmegen, The Netherlands; Texas A&M University, College Station, TX, United States; Avon and Wiltshire Partnership NHS Mental Health Trust, Bristol, United Kingdom; NIHR Biomedical Research Centre, University of Bristol, Bristol, United Kingdom

## Abstract

Common genetic variation has been associated with multiple symptoms in Autism Spectrum Disorder (ASD). However, our knowledge of shared genetic factor structures contributing to this highly heterogeneous neurodevelopmental condition is limited. Here, we developed a structural equation modelling framework to directly model genome-wide covariance across core and non-core ASD phenotypes, studying autistic individuals of European descent using a case-only design. We identified three independent genetic factors most strongly linked to language/cognition, behaviour and motor development, respectively, when studying a population-representative sample (N=5,331). These analyses revealed novel associations. For example, developmental delay in acquiring personal-social skills was inversely related to language, while developmental motor delay was linked to self-injurious behaviour. We largely confirmed the three-factorial structure in independent ASD-simplex families (N=1,946), but uncovered simplex-specific genetic overlap between behaviour and language phenotypes. Thus, the common genetic architecture in ASD is multi-dimensional and contributes, in combination with ascertainment-specific patterns, to phenotypic heterogeneity.

## Introduction

Autism spectrum disorder (ASD) is a complex neurodevelopmental condition with considerable phenotypic and genetic heterogeneity (1,2). Core phenotypes in ASD implicate difficulties in social interaction and communication, as well as restricted, repetitive behavioural patterns and sensory abnormalities (3). However, the phenotypic presentation is broad and variable. More than 70% of individuals with ASD are diagnosed with co-occurring conditions (henceforth referred to as ASD phenotypic spectrum) (4), and individuals differ in phenotypic presentation, especially cognitive functioning (2,4). At the genetic level, additive genetic effects of rare and common genetic factors contribute to ASD liability in a sex-specific manner (1,5–10). Common variation explains most genetic variance in ASD, accounting for 12 to 65% of liability (1,5,11). However, even common genetic variation is highly heterogenous in ASD (5,6,8), and differences in underlying shared genetic factors are only partially understood.

Depending on an individual’s genetic architecture, common variants act through partially distinct aetiological mechanisms (6). For example, autistic individuals with intellectual disability (ID), compared to those without, carry a higher rate of contributing *de novo* variants (6) and show qualitative differences in their common genetic architecture (5). In addition, polygenic scores (PGS) for different disorders, aggregating common risk alleles, show distinct association profiles with phenotypic factor structures in groups comprising only autistic individuals (8,12). Thus, also common variation may present genetic factor structures linking phenotypic domains, although the number of factors and their nature is unknown. Furthermore, the genetic architecture of ASD is distinctly different in multiplex families with multiple affected family members, compared to simplex families with only one affected child (13). ASD liability in simplex families is considerably more often related to *de novo* mutations (11,14). Therefore, also common genetic factor structures may differ between exclusively simplex and population-representative ASD architectures, where latter contain both simplex and multiplex families.

This study applies genetic-relationship-matrix (GRM) structural equation modelling (GRM-SEM) techniques to identify and characterise shared genetic factor structures in autistic individuals from large ASD cohorts adopting a case-only design (**Figure 1**). GRM-SEM estimates multivariate common genetic architectures (15), as captured by GRMs derived from direct genotyping data (15,16), by directly fitting structural models to genetic and residual variation using a maximum likelihood (ML) approach (15). Consequently, models can be compared with and optimised against a saturated model, i.e. a model with a perfect fit (15). Here, we introduce a data-driven version of GRM-SEM (**Figure 1B**) that minimises the computational burden of identifying the best-fitting multi-dimensional model. We predict the number of genetic factors and their structure from genetic trait covariance, as estimated with a saturated GRM-SEM model, using principal component analysis (PCA) and exploratory factor analysis (EFA) techniques. Given the estimated nature of these genetic data, conducted analyses are approximations only, henceforth referred to as genetic PCA and genetic EFA, respectively. We use genetic PCA and EFA information to identify a multi-dimensional GRM-SEM model by providing starting values and parameter constraints.

**Figure 1.**
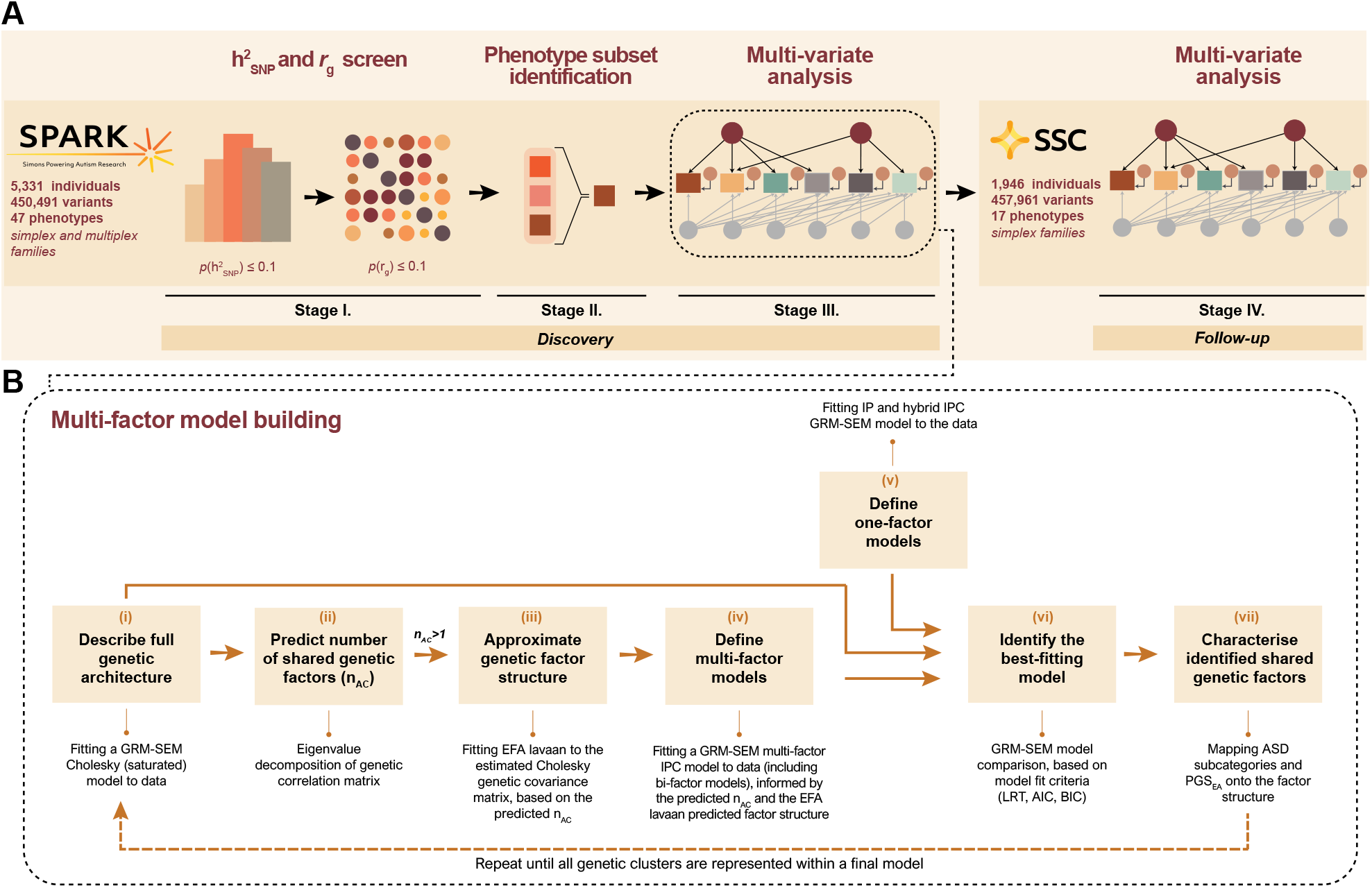
Workflow of the study. **(A)** Multivariate discovery analyses were carried out in the Simons Powering Autism Research (SPARK) sample (Stages I-III) and the best-fitting final SPARK multi-factor model was followed-up in the Simons Simplex Collection (SSC, Stage IV). **(B)** Data-driven approach to model genomic covariance with genetic-relationship-matrix structural equation modelling (GRM-SEM). We fitted (i) a saturated GRM-SEM (Cholesky) model to describe the genetic architecture. Based on this information, we (ii) predicted the number of shared genetic factors (n_AC_) across phenotypes through eigenvalue decomposition of Cholesky-derived genetic correlations. If n_AC_ >1, we (iii) approximated the underlying genetic factor structure through exploratory factor analysis (EFA) of Cholesky-derived genetic trait covariance. We used this information on genetic factor structures from (ii) and (iii) to fit (iv) multi-factor Independent Pathway/Cholesky (IPC) models, including bi-factor models (to confirm the independence of shared genetic factors). For comparison only, we fitted (v) one-factor Independent Pathway (IP) and IPC models, analogous to twin analyses. We compared (vi) the model fit of multi-factor models to one-factor models and the saturated model to identify the best-fitting model. This multistep approach was repeated until all phenotype subsets were combined into a final model. Eventually, we (vii) characterised the factor structure of the final best-fitting model by mapping it to a clinical reference (DSM-IV-based ASD subcategories) and to the polygenic score for educational attainment (PGS_EA_), enhancing the interpretability of predicted factor structures.

We implement this data-driven modelling strategy into a multi-stage research design to examine the genetic architecture of ASD across a broad range of related phenotypes and comorbidities, studying the most well-characterised ASD cohorts to date. As part of discovery analyses, we investigate ASD core and non-core phenotypes for 5,331 European descent individuals with ASD from the Simons Foundation Powering Autism Research for Knowledge (SPARK) sample (17) (**Supplementary Table 1, Supplementary Figure 1, Supplementary Methods 1**). Recruited across the United States (US), SPARK is a population-representative ASD sample including individuals from simplex or multiplex families (17). We follow up our results on 1,946 autistic individuals from simplex-only families of the Simons Simplex Collection (SSC) sample (18) (**Supplementary Table 2, Supplementary Figure 2, Supplementary Methods 2**). Here, we report and contrast comprehensive multivariate genetic models estimated in SPARK and the SSC, empowering new insights into the heterogeneous phenotypic spectrum of ASD across samples representing different ascertainment schemes.

## RESULTS

### Multi-dimensional genetic analyses in population-representative ASD

A challenge in identifying the genetic architecture of ASD core and non-core phenotypes is the selection of measures for model building. We, therefore, conducted discovery analyses in the population-representative SPARK sample across multiple stages (**Figure 1**). During stage I, we screened for phenotypes that are likely to have some genetic contributions (h^2^_SNP_, *p*≤0.1, **Figure 2A**), using Genomic Restricted Maximum Likelihood (GREML) embedded in GCTA software (19,20), to ensure the convergence of GRM-SEM models. Note that h^2^_SNP_ estimates in this study reflect phenotypic heterogeneity among autistic individuals that can be accounted for by common genetic variation. We retained 17 phenotypes from an initial set of 47 phenotypes, disorders and developmental milestones (**Figure 2A**). Captured domains included language/cognition, general behaviour, developmental, motor and repetitive behavioural features. Social and affective phenotypes were not taken forward to the next stage of analysis due to a lack of evidence for h^2^_SNP_ (**Supplementary Figure 3**). Next, we screened for phenotype combinations that are possibly sharing common genetic variation (*r*_g_ *p*≤0.1, **Figure 2B**) to enable the identification of overarching genetic factors.

**Figure 2.**
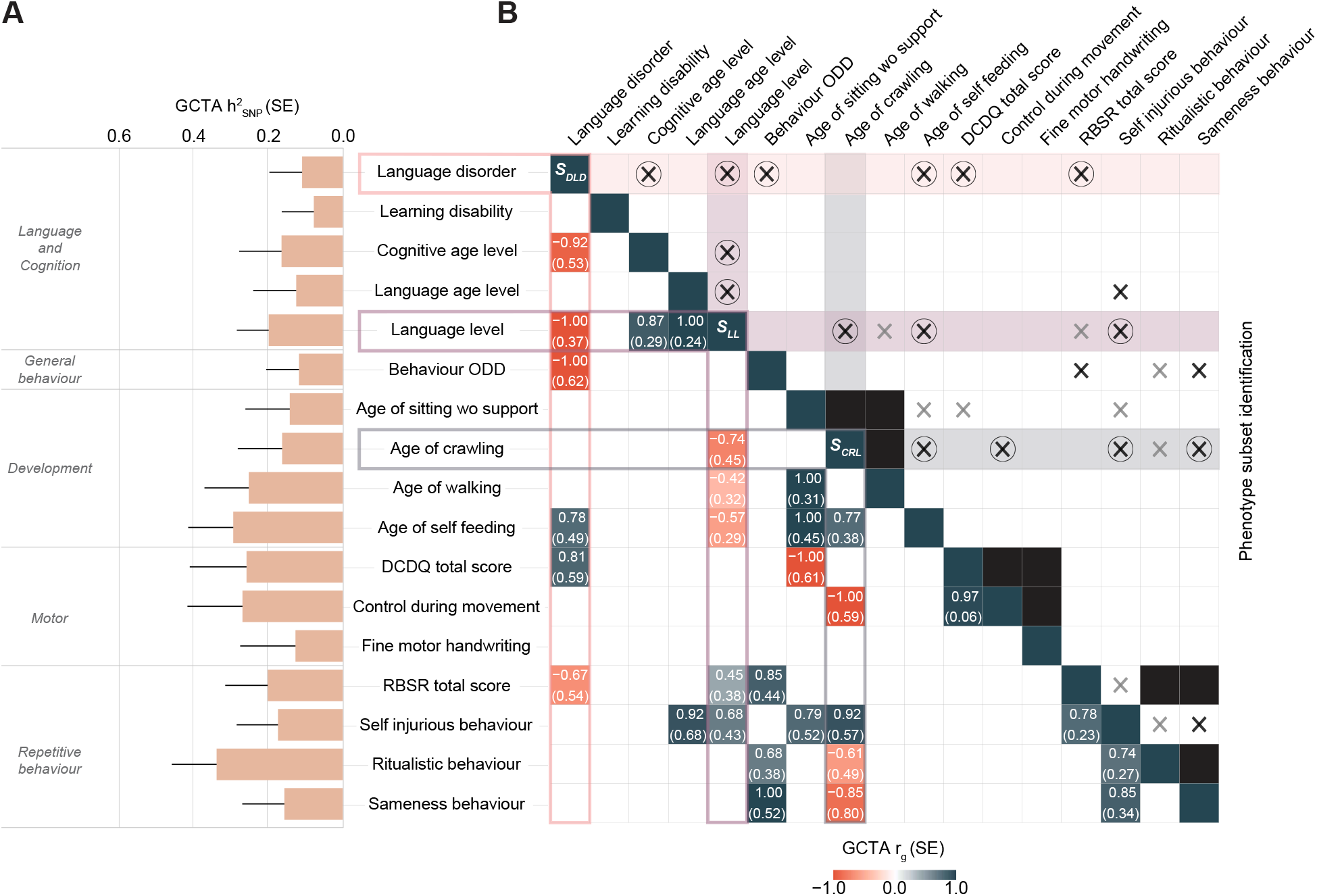
Screen for heritable and genetically interrelated phenotypes in SPARK. **(A)** Heritability (h^2^_SNP_) of continuous and categorical ASD phenotypes (p≤0.1) as estimated by GCTA. A complete figure of all analysed phenotypes is shown in Supplementary Figure 3. The error bars represent standard errors. Estimates were based on transformed scores: deviance residuals (for categorical phenotypes) or rank-transformed residuals (for continuous phenotypes). **(B)** The lower triangle shows the genetic correlation screen (r_g_) across ASD phenotypes as shown in (A), passing p(r_g_)≤0.1, as estimated with GCTA. A complete figure of all correlations is shown in Supplementary Figure 4. The upper triangle shows the selected phenotype subsets that, together, comprehensively capture the genetic correlations (lower triangle) across studied phenotypes. Each phenotypic subset has a ‘node’ phenotype: S_DLD_ (language disorder), S_LL_ (language level) and S_CRL_ (age of crawling). Phenotypes within a subset are directly genetically correlated with the ‘node’ phenotype (p≤0.1). The black boxes symbolise proxy phenotypes, as identified within uni-factorial GRM-SEM (r_g_=1, Supplementary Figure 5). Circled ‘x’ within shaded boxes indicate the phenotypes that are included in each subset and were directly modelled with GRM-SEM. A black ‘x’ indicates directly estimated and a grey ‘x’ indirectly (proxied) genetic relationships. Phenotypes were adjusted for covariates and transformed into either rank-transformed residuals (continuous measures) or deviance residuals (categorical measures). *Abbreviations*: DCDQ (Developmental Coordination Disorder Questionnaire), GCTA (Genome-wide Complex Trait Analysis), GRM-SEM (Genetic Relationship Matrix Structural Equation Modelling), ODD (oppositional defiant disorder), RBSR (Repetitive Behaviour Scale-Revised).

Within stage II, we selected phenotypic subsets that jointly captured estimated genetic correlations from stage I, based on an enumeration of phenotype combinations, with the aim to successively construct a comprehensive GRM-SEM model (**Figure 2B, Supplementary Figure 4, Supplementary Table 3, Supplementary Note 1**). Building a model from smaller phenotypic subsets ensures the robustness of identified structures and reduces the computational burden. The most extensively genetically linked phenotypic subsets were related to language disorder (including developmental language disorder/delay, S_DLD_), language level (S_LL_) and age of crawling (S_CRL_), respectively (**Figure 2B**). To control for measurement collinearity that can affect model convergence, we searched, in addition, for genetically correlated scales within a questionnaire using uni-dimensional GRM-SEM (**Supplementary Note 1, Supplementary Figure 5**). Where item scales of the same instrument were genetically similar (GRM-SEM *r*_g_=1), we retained a single representative measure (or proxy) only (**Figure 2B, Supplementary Note 1, Supplementary Figure 5**).

As part of stage III, we aimed to identify the best-fitting multi-dimensional GRM-SEM models for the selected phenotypic subsets and, eventually, a combined set of measures, S_ALL_ (**Supplementary Note 2**). For this, we fitted a series of GRM-SEM saturated (Cholesky) models, genetic PCA eigenvalue decompositions, genetic EFA models, and, finally, GRM-SEM multi-dimensional and bi-factor models (**Figure 1, Table 1, Supplementary Table 4, Methods, Supplementary Note 3**). For the latter two model types, we adopted a hybrid Independent Pathway / Cholesky (IPC) design that has shown a superior fit in previous analyses (16), as the residual part of the data is always fitted to a saturated (Cholesky) model (**Methods**).

**Table 1.** Model fit comparison.

| Model | Type | log-likelihood | N<sub>par</sub> | AIC | BIC | LRT<sub>Cholesky</sub> |  | LRT<sub>Bi-factor</sub> |  |
| --- | --- | --- | --- | --- | --- | --- | --- | --- | --- |
| | | | | | | $\Delta\chi^2(\Delta\text{df})$ | p | $\Delta\chi^2(\Delta\text{df})$ | p |
| SPARK |  |  |  |  |  |  |  |  |  |
| S<sub>DLD</sub> model: N<sub>traits</sub>=7, N<sub>ind</sub>=5279 |  |  |  |  |  |  |  |  |  |
| Cholesky | saturated | -13343.57 | 56 | 26799.13 | 27167.14 | - |  | - |  |
| Bi-factor | two-factor | -13345.64 | 46 | 26783.28 | 27085.56 | 4.14(10) | 0.94 | - |  |
| IPC best-fit | two-factor | -13345.66 | 40 | **26771.33** | **27034.19** | 4.19(16) | 1.00 | 0.05(6) | 1.00 |
| S<sub>LL</sub> model: N<sub>traits</sub>=7, N<sub>ind</sub>=5279 |  |  |  |  |  |  |  |  |  |
| Cholesky | saturated | -12524.86 | 56 | 25161.72 | 25529.72 | - |  | - |  |
| Bi-factor | two-factor | -12526.61 | 46 | 25145.23 | 25447.51 | 3.51(10) | 0.97 | - |  |
| IPC best-fit | two-factor | -12527.13 | 41 | **25136.27** | **25405.70** | 4.55(15) | 1.00 | 1.04(5) | 0.96 |
| S<sub>ALL</sub> (S<sub>DLD</sub>, S<sub>LL</sub> and S<sub>CRL</sub>) model: N<sub>traits</sub>=8, N<sub>ind</sub>=5279 |  |  |  |  |  |  |  |  |  |
| Cholesky | saturated | -15248.61 | 72 | 30641.23 | 31114.37 | - |  | - |  |
| Bi-factor | three-factor | -15249.97 | 62 | 30623.94 | 31031.37 | 2.71(10) | 0.99 | - |  |
| IPC best-fit | three-factor | -15250.96 | 53 | **30607.92** | **30956.21** | 4.69(19) | 1.00 | 1.98(9) | 0.99 |
| SSC |  |  |  |  |  |  |  |  |  |
| S<sub>ALL</sub> model: N<sub>traits</sub>=8, N<sub>ind</sub>=1940 |  |  |  |  |  |  |  |  |  |
| Cholesky | saturated | -6342.50 | 72 | 12828.99 | 13230.07 | - |  | - |  |
| Bi-factor | three-factor | -6342.59 | 63 | 12811.18 | 13162.12 | 0.19(9) | 1.00 | - |  |
| IPC best-fit | three-factor | -6342.60 | 53 | **12791.19** | **13086.43** | 0.20(19) | 1.00 | 0.01(10) | 1.00 |

**Abbreviations:** AIC (Akaike information criterion); BIC (Bayesian information criterion); IPC (Hybrid Independent Pathway (genetic part) / Cholesky (residual part) model); N<sub>par</sub> (number of parameters), S<sub>CRL</sub> (age of crawling subset), S<sub>DLD</sub> (language disorder subset), S<sub>LL</sub> (language level subset), and S<sub>SSC</sub> (follow-up subset).

For all phenotypic sets with unambiguously identified numbers of genetic factors (S_DLD_, S_LL_ and S_ALL_), genomic structures were predicted with genetic EFA. Here, we fitted orthogonal (varimax) genetic EFA throughout, given modest genetic factor correlations (**Supplementary Note 2, Supplementary Tables 5-6**). Based on the identified best-fitting GRM-SEM models for S_DLD_ and S_LL_ (**Table 1, Supplementary Table 4, Figure 3A-F, Supplementary Tables 7-8, Supplementary Figure 6-7**), we, eventually, added phenotypes to a combined set S_ALL_. To reduce the computational burden, we selected S_DLD_ and S_LL_ measures with both the most substantial factor (|λ|>0.3) and cross-factor loadings (|λ|>0.1), representing all phenotypic domains (**Supplementary Table 3, Supplementary Note 2**). For the S_CRL_ subset, eigenvalue decomposition did not reveal an exact factor dimension (**Supplementary Figure 8**) and, therefore, the entire set was added to S_ALL_ (**Supplementary Note 2**). Once S_ALL_ building was completed, we repeated the modelling process as described above.

**Figure 3.**
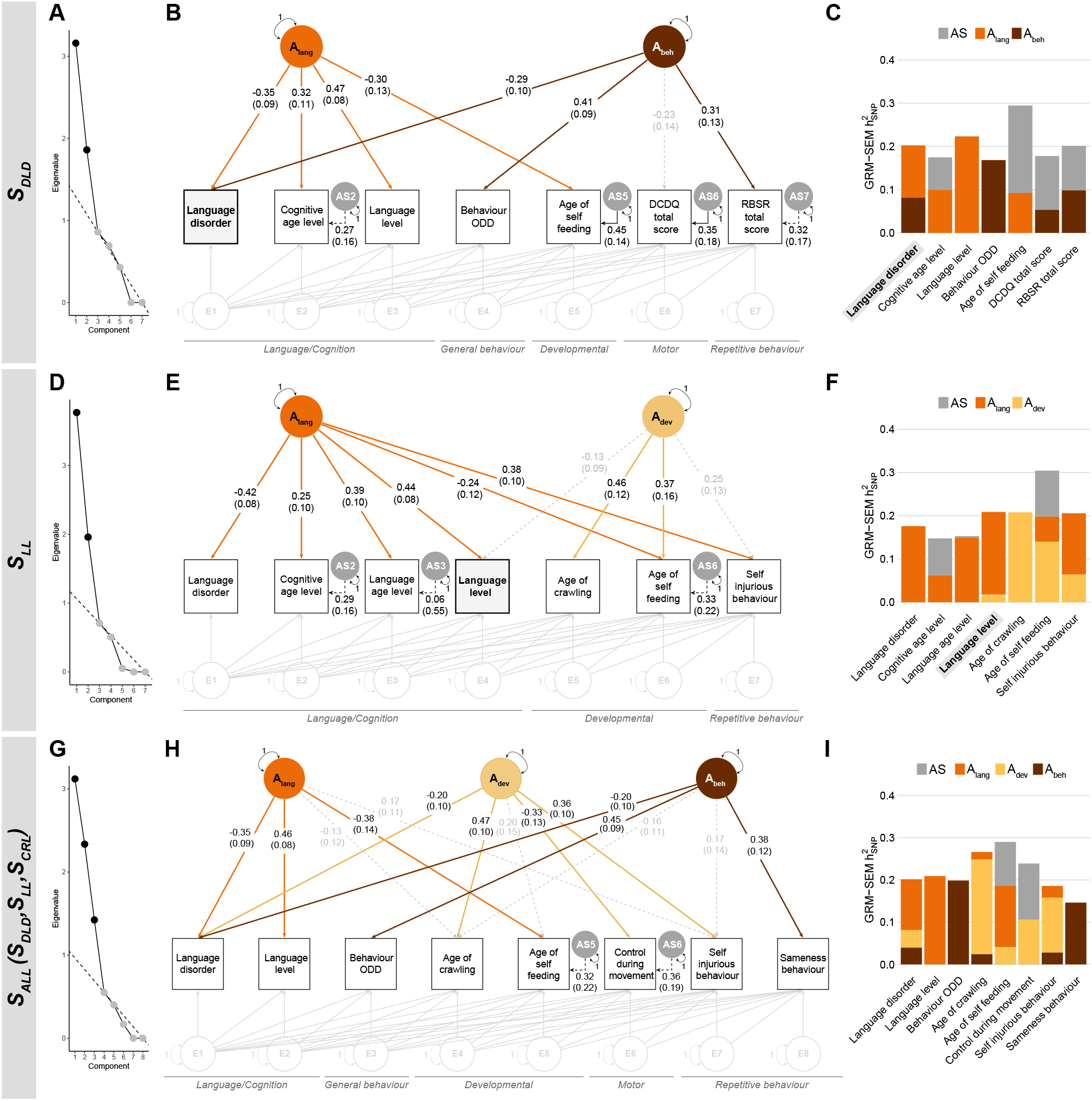
Multi-factor GRM-SEM models in SPARK. **(A)** Scree plot, **(B)** path diagram and **(C)** standardised genetic variance (GRM-SEM h^2^_SNP_) plot of the best-fitting GRM-SEM IPC model for the language disorder (S_DLD_) set. **(D)** Scree plot, **(E)** path diagram and **(F)** standardised genetic variance (GRM-SEM h^2^_SNP_) plot of the best-fitting GRM-SEM model for the language level (S_LL_) set. **(G)** Scree plot, **(H)** path diagram and **(I)** standardised genetic variance (GRM-SEM h^2^_SNP_) plot of the best-fitting GRM-SEM model for the combined (S_ALL_: S_DLD_, S_LL_ and S_CRL_ set) set. **(A**,**D**,**G)** Scree plots are based on the eigenvalue decomposition of genetic correlations derived from a GRM-SEM Cholesky model, depicting the number of estimated shared genetic factors (in black) according to the optimal coordinate criterion. The dashed line indicates the “scree” of the plot (grey). **(B**,**E**,**H)** Observed measures are represented by squares and latent variables by circles (A: shared genetic factor, AS: specific genetic factor, E: residual factor). Dotted and solid single-headed arrows (factor loadings) define relationships between variables with *p*>0.05 and *p*≤0.05, respectively. The genetic part of the model has been modelled using an Independent Pathway model, and the residual part using a Cholesky model (grey). (**C**,**F**,**I)** SEs for GRM-SEM h^2^_SNP_ contributions have been omitted for clarity. Note that no GRM-SEM model was fitted to the third S_CRL_ (age of crawling) subset, as the number of genetic factors could not be unambiguously predicted by the optimal coordinate criterion. *Abbreviations*: A_lang_ (Genetic language factor), A_dev_ (Genetic developmental-delay factor), A_beh_ (Genetic behavioural-problems factor), DCDQ (Developmental Coordination Disorder Questionnaire), h^2^_SNP_ (Single nucleotide polymorphism-based heritability), IPC (Independent Pathway-Cholesky GRM-SEM model), ODD (Oppositional Defiant Disorder), RBSR (Repetitive Behaviours Scale-Revised).

For each modelled phenotype set, S_DLD_ (**Figure 3A-C, Supplementary Table 7, Supplementary Figure 6**), S_LL_ (**Figure 3D-F, Supplementary Table 8, Supplementary Figure 7**) and S_ALL_ (**Figure 3G-I, Supplementary Table 9, Supplementary Figure 9**), a multi-dimensional IPC model fitted the data best (**Table 1, Supplementary Table 4**), matching predicted eigenvalues. Model comparisons were based on Akaike and Bayesian information criteria (AIC and BIC), and likelihood ratio tests (LRTs), and the fit of all identified models was highly comparable to a saturated model (*p*_LRT_=1; **Table 1, Supplementary Table 4**). Across subsets (S_DLD_ and S_LL_) and the combined set (S_ALL_), we found stable genetic dimensions (**Figure 3, Supplementary Tables 7-8, Supplementary Figures 6-7,9, Supplementary Note 2**) demonstrating the robustness of underlying genetic structures.

The combined set (S_ALL_), comprised two language phenotypes (language disorder, language level), oppositional defiant disorder (ODD) as a form of general behavioural problems, two developmental milestones (age of self-feeding, age of crawling), control during movement as a proxy for Developmental Coordination Disorder Questionnaire (DCDQ) motor scores and two Repetitive Behaviour Scale-Revised (RBSR) behaviour scores (self-injurious behaviour, sameness behaviour) (**Figure 3H-I**). A three-factor IPC model fitted the data best (**Figure 3G-I, Table 1, Supplementary Table 4**). The three identified factors captured most strongly language (A_lang_), developmental delay (A_dev_) and behavioural problems (A_beh_) (**Figure 3H, Supplementary Figure 9, Supplementary Table 9**), consistent with S_DLD_ and S_LL_ models (**Figure 3B**,**3E**). To explore the factor structure, we focused on standardised genetic factor loadings with an explanatory value of |λ|≥0.3 (21), accounting for ∼10% phenotypic or liability variation, as well as the factorial coheritability (f^2^_g_), i.e. the fraction of h^2^_SNP_ that is explained by a factor.

The first genetic factor captured better language performance, A_lang_ (**Figure 3H**) and was most strongly related to better language level (λ_lang_= 0.46, SE=0.08), lower liability to language disorder (λ_lang_=-0.35, SE=0.09) and earlier age of self-feeding (λ_lang_=-0.38, SE=0.14). Across phenotypes, this factor accounted for at least half of the trait h^2^_SNP_ estimates (f^2^_g_, 0.50-1.00, **Supplementary Table 9**). Notably, this factor also uncovered inverse correlations between children’s language ability (e.g. language level) and the age of self-feeding (GRM-SEM *r*_g_=-0.71, SE=0.25, **Figure 4A, Supplementary Figure 9**).

**Figure 4.**
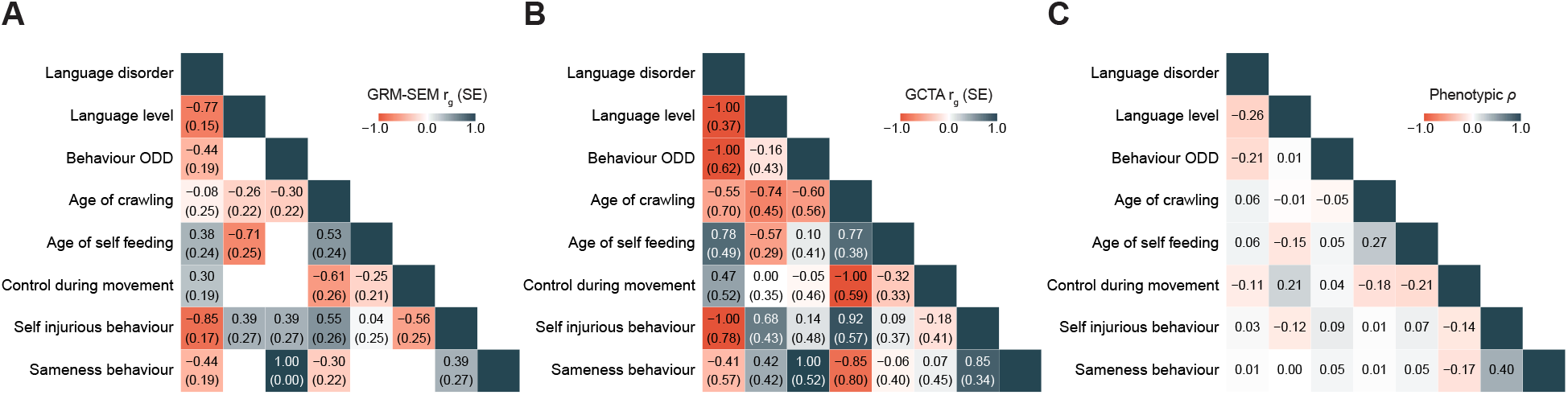
Correlations for the combined (S_ALL_) phenotypic subset. Figure shows **(A)** GRM-SEM genetic correlations, **(B)** GCTA genetic correlations and **(C)** Spearman phenotypic correlations. All correlations are based on transformed measures. *Abbreviations*: GCTA (Genome-wide Complex Trait Analysis), GRM-SEM (Genetic Relationship Matrix Structural Equation Modelling), ODD (oppositional defiant disorder).

The second genetic factor, A_dev_, reflecting developmental delay, captured a later age of crawling (λ_dev_=0.47, SE=0.10), less motor control (DCDQ control during movement, λ_dev_=-0.33, SE=0.13) and more RBSR self-injurious behaviour (λ_dev_=0.36, SE=0.10), explaining a considerable proportion of genetic variance (f^2^_g_ =0.44-0.84, **Supplementary Table 9**). The third genetic factor, A_beh_, was linked to behaviour problems, almost fully explaining the h^2^_SNP_ of each trait (f^2^_g_ =1), including RBSR sameness behaviour (λ_beh_ =0.38, SE=0.12) and liability to ODD (λ_beh_=0.45, SE=0.09).

Identified genetic factors largely matched corresponding phenotypic dimensions. Each phenotype had a single meaningful factor loading (|λ|>0.3) for one factor only (21). However, for liability to language disorder, cross-loadings (*p*<0.05) with all three factors were detected (λ_lang_=-0.35, SE=0.09; λ_dev_=-0.20, SE=0.10; λ_beh_=-0.20, SE=0.10), indicating genetic heterogeneity. Given the broad phenotypic definition of developmental language delay and disorder, genetic links across independent genetic dimensions may arise due to multiple underlying conditions (22).

Further heterogeneity in genetic links was uncovered for self-injurious behaviour. Despite overall stability in factor structures, RBSR self-injurious behaviour, depending on the studied context, was either genetically related to the language A_lang_ factor (λ_lang_=0.38, SE=0.10, **Figure 3E**, S_LL_ model) or the developmental-delay-related A_dev_ factor loading (λ_dev_=0.36, SE=0.10, **Figure 3H**, S_ALL_ model). Genetic cross-loadings with two independent common dimensions suggest distinct genetic aetiologies (22), matching different forms of self-injurious behaviour in ASD. While some forms involve stereotyped and repetitive behaviour, co-morbid with ID (23), others show neurotypical patterns (24,25) that facilitate cognitive regulation such as the release of ‘high pressure’ emotions (24,25). In contrast, there was little evidence for genetic links of self-injurious behaviour with the behavioural-problem’s factor (**Figure 3E**,**3H**), matching previously reported distinct phenotypic factor structures (8). Thus, self-injurious actions may, at least partially, be aetiologically distinct from other forms of repetitive behaviour.

Next, we confirmed the independence of predicted genetic factors by conducting bi-factor models, each showing a similar fit (*p*_LRT_≥0.94, **Table 1, Supplementary Figures 10-12**). In addition, we corroborated predicted r_g_ (**Figure 4**) and h^2^_SNP_ patterns (**Supplementary Figure 13**), derived from the best-fitting GRM-SEM model for the S_ALL_ set, through comparisons with corresponding GREML analyses. We observed consistent findings throughout, based on 95% CIs, demonstrating that genetic dimensions and structure of multivariate genetic architectures can be accurately predicted by genetic PCA and EFA analyses (**Figure 3A**,**3D**,**3G**), analogous to methodologies developed for summary statistics (26).

Eventually, to enhance the interpretability of identified genetic structures, we mapped ASD subcategory information and PGS for educational attainment (EA) onto the model structure of the S_ALL_ model in SPARK, while preserving the model fit (**Figure 3H** versus **Figure 5A,5D**, **Supplementary Table 4**). ASD subcategory information (DSM-IV-based) can provide a clinical reference guiding the interpretation of identified cognitive genetic dimensions, here capturing genetic liability to Asperger, a form of autism without significant impairments in language and cognitive development (27). In contrast, PGS_EA_ presents a genetic correlate of cognitive functioning (28), but also socio-economic status, including health and longevity (29). Here, once mapped, liability to Asperger was genetically linked to the language genetic factor (**Figure 5A**, λ_lang_=0.36, SE=0.15). Genetic correlations between liability to Asperger and language level (**Figure 5C**, GRM-SEM *r*_g_=0.90, SE=0.19) were positive, consistent with the absence of language problems in this ASD subcategory (3). In contrast, PGS_EA_ were inversely associated with the behavioural problem factor (**Figure 5D**, λ_beh_=-0.16, SE=0.06), conditional on the language/cognitive dimension. Consistently, genetic correlations of PGS_EA_ with behavioural measures such as sameness behaviour were inverse (**Figure 5F**, GRM-SEM *r*_g_=-0.16, SE=0.06), strengthening support for previously reported links with repetitive behaviour (9).

**Figure 5.**
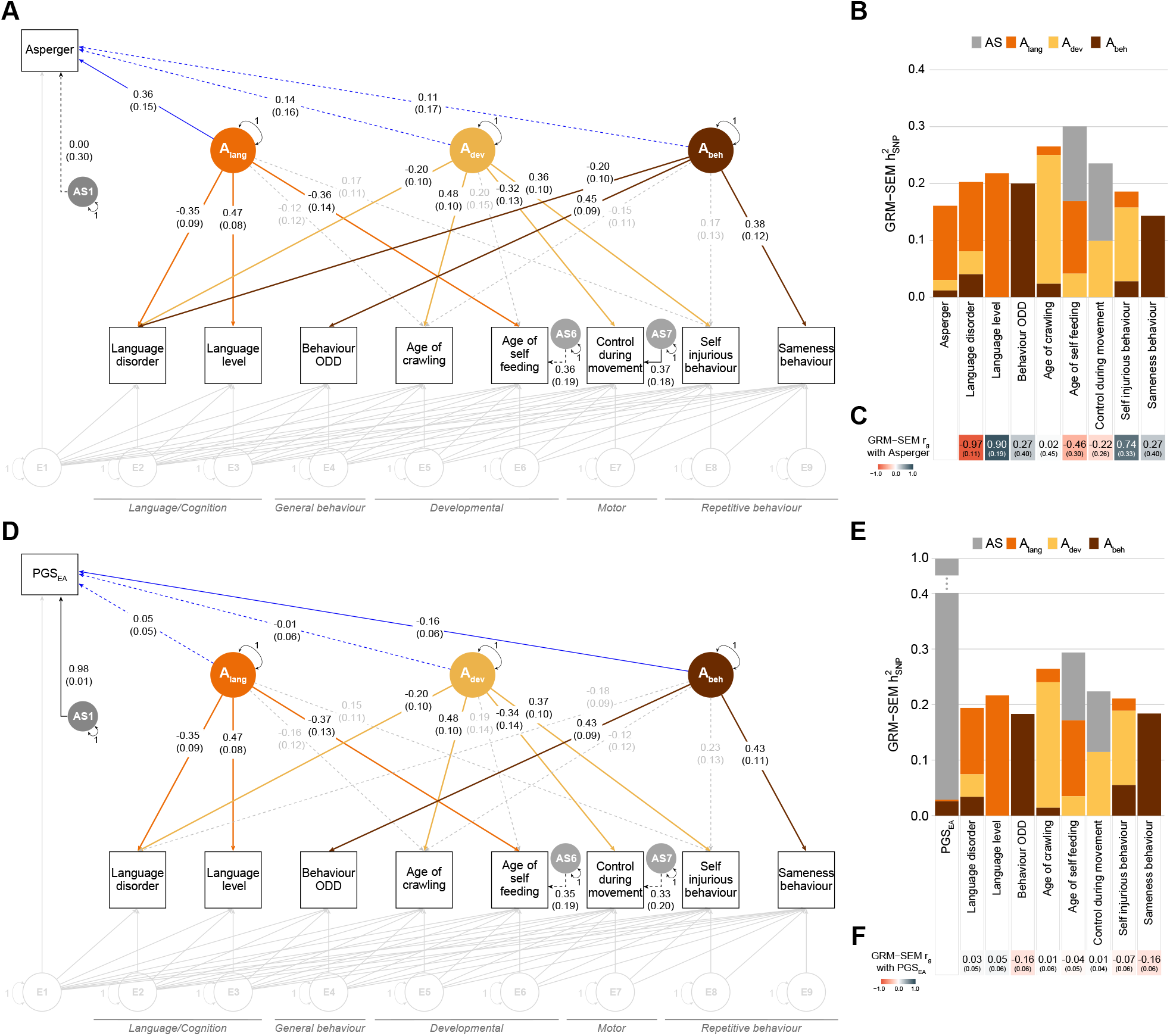
ASD subcategory mapping of the multi-factor GRM-SEM model for the combined (S_ALL_) set in SPARK. **(A)** Path diagram of an extended GRM-SEM IPC model mapping liability to Asperger (reference: Asperger against other ASD subcategories) onto the model structure of the best-fitting (S_ALL_, Figure 3H) SPARK model. **(B)** Corresponding standardised genetic variance (GRM-SEM h^2^_SNP_) plot. SEs for GRM-SEM h^2^_SNP_ contributions have been omitted for clarity. **(C)** Genetic correlations with liability to Asperger. **(D)** Path diagram of an extended GRM-SEM IPC model mapping the polygenic score for educational attainment (PGS_EA_) onto the model structure of the best-fitting (S_ALL_, Figure 3H) SPARK model. **(E)** Corresponding standardised genetic variance (GRM-SEM h^2^_SNP_) plot. SEs for GRM-SEM h^2^_SNP_ contributions have been omitted for clarity. **(F)** Genetic correlations with the PGS_EA_. **(A**,**D)** Observed measures are represented by squares and latent variables by circles (A_lang_/A_dev_/A_beh_: shared genetic factor, AS: specific genetic factor, E: residual factor). Dotted and solid single-headed arrows (factor loadings) define relationships between variables with *p*>0.05 and *p*≤0.05, respectively. The genetic part of the model has been modelled using an Independent Pathway model, and the residual part using a Cholesky model (grey). *Abbreviations:* A_lang_ (Genetic language factor), A_dev_ (Genetic developmental-delay factor), A_beh_ (Genetic behavioural-problems factor), DCDQ (Developmental Coordination Disorder Questionnaire), h^2^_SNP_ (Single nucleotide polymorphism-based heritability), IPC (Independent Pathway-Cholesky GRM-SEM model), ODD (Oppositional Defiant Disorder), RBSR (Repetitive Behaviours Scale-Revised), r_g_ (genetic correlation).

Note that low sample numbers and/or low h^2^_SNP_ of ASD liability prevented a more comprehensive modelling (**Supplementary Figure 14**).

### Multi-dimensional genetic analyses in simplex ASD

Within stage IV, we attempted to reproduce the best-fitting GRM-SEM model identified in the population-representative SPARK sample (S_ALL_) by studying ASD individuals from SSC simplex families. Matching SSC phenotypes showed little evidence for h^2^_SNP_ (**Supplementary Figure 15**), consistent with the smaller sample size. Both motor (DCDQ scores) and self-injurious behaviour (RBSR) scores had to be excluded from follow-up due to near-zero h^2^_SNP_ point estimates. These two measures were replaced with further language/cognition and developmental phenotypes to allow for an empirical identification of three genetic dimensions. The final phenotype subset (S_SSC_) reflected phenotypes studied in SPARK: three language/cognition measures (language disorder, language age level, language level), general behaviour (ODD), three developmental milestones (age of crawling, age of self-feeding, age of walking), and the RBSR repetitive behaviour score (sameness behaviour).

As in SPARK, a three-factor model (**Figure 6**, **Supplementary Table 10**) fitted the data best (**Table 1**, **Supplementary Table 4**), matching predicted eigenvalues. The first genetic factor (A_F1_) accounted for variation in language age level (λ_F1_=0.33,SE=0.14; f^2^_g_=0.21,SE=0.16) and age of self-feeding (λ_F1_=-0.46,SE=0.19; f^2^_g_ =1.00,SE<0.01), corresponding to the A_lang_ factor structure in SPARK (**Figure 3B,3E,3H**, **Figure 6B**). Note, within SPARK, language level (i.e. an individual’s everyday language skills) and language age level (i.e. an individual’s spoken language for their age level) are strongly correlated (GCTA *r*_g_=1.00,SE=0.24) and showed, when modelled together, similar association patterns (e.g. S_LL_ model, **Supplementary Figure 7**). The second genetic factor (A_F2_) described variation across developmental-delay-related phenotypes, with the strongest factor loading for age of walking (λ_F2_=0.62,SE=0.14; f^2^_g_ =0.93,SE=0.22), comparable to the A_dev_ factor structure in SPARK (**Figure 3E,3H**, **Figure 6B**). The third genetic factor (A_F3_) (**Figure 6B**) explained shared genetic variation (f^2^_g_ =0.75-1.00, **Supplementary Table 10**) across language/cognition and repetitive (RBSR sameness) behaviour. The strongest factor loadings were observed for language age level (λ_F3_=0.61,SE=0.10), language disorder (λ_F3_=-0.51,SE=0.11), language level (λ_F3_=0.37,SE=0.07), but also RBSR sameness behaviour (λ_F3_=0.51, SE=0.12). This cross-trait genetic dimension in the SSC captured strong positive genetic correlations between language and repetitive behaviour (e.g. language level, RBSR sameness behaviour: GRM-SEM *r*_g_=0.97, SE=0.07, **Figure 6D**) that were absent in SPARK (language level, RBSR sameness behaviour: GRM-SEM *r*_g_=0, **Supplementary Figure 9**).

**Figure 6.**
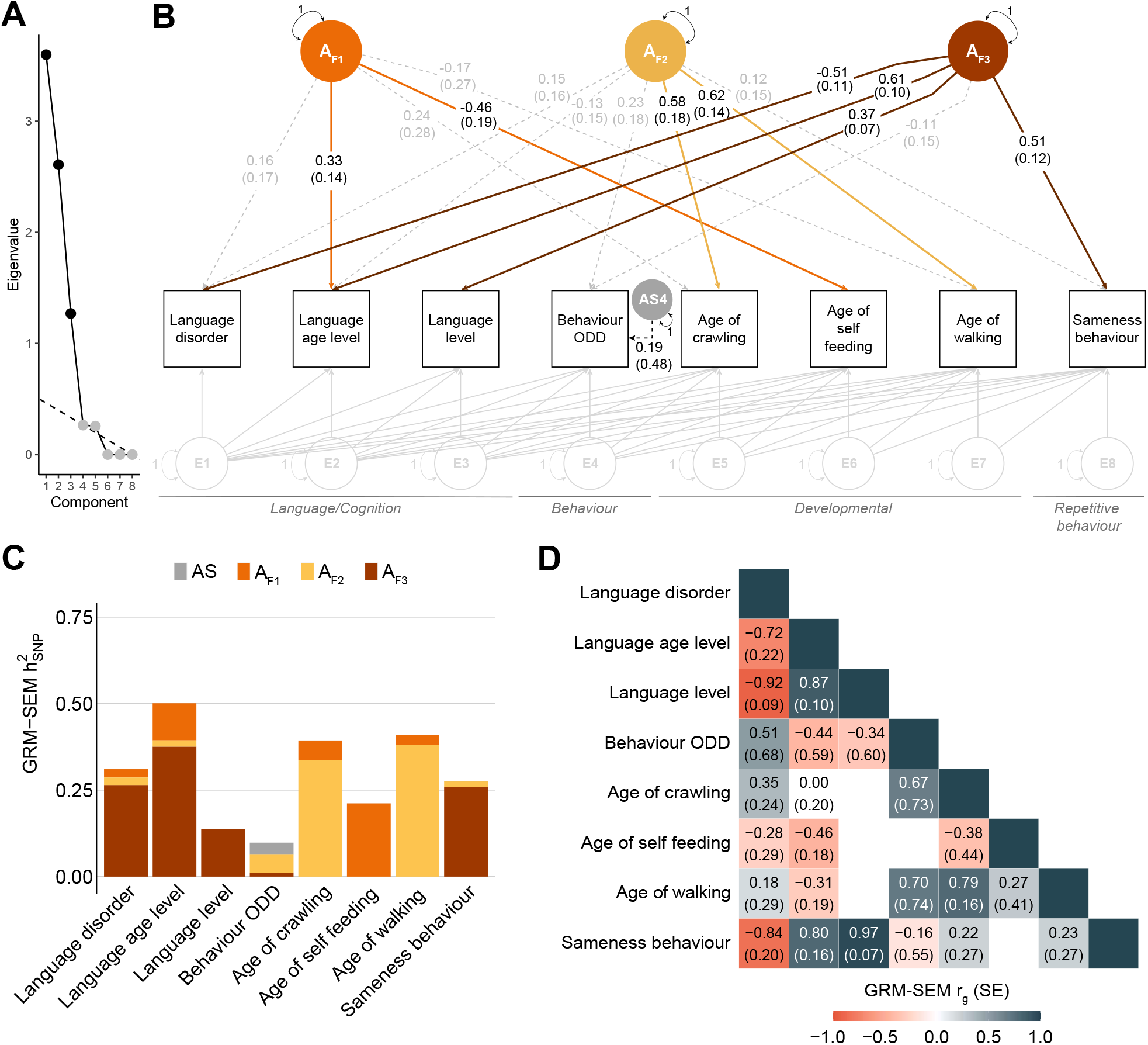
Follow-up multi-factor GRM-SEM model in the SSC (S_SSC_). **(A)** Scree plot based on the eigenvalue decomposition of genetic correlations derived from a GRM-SEM Cholesky model, depicting the number of estimated shared genetic factors (in black) according to an optimal coordinate criterion. The dashed line indicates the “scree” of the plot (grey). **(B)** Path diagram depicting the best-fitting multi-dimensional GRM-SEM IPC model based on largely comparable phenotypes as studied in SPARK. Observed measures are represented by squares and latent variables by circles (A: shared genetic factor, AS: specific genetic factor, E: residual factor). Dotted and solid single-headed arrows (factor loadings) define relationships between variables with p>0.05 and p≤0.05, respectively. The genetic part of the model has been modelled using an Independent Pathway model, and the residual part using a Cholesky model (grey). **(C)** Corresponding standardised genetic variance (GRM-SEM h^2^_SNP_) plot. SEs for GRM-SEM h^2^_SNP_ contributions have been omitted for clarity. **(D)** Corresponding correlogram of genetic correlations. Numeric values for genetic correlations that are not predicted by the genetic model structure were omitted. *Abbreviations*: A_F1,2,3_ (Genetic factor 1,2,3), h^2^_SNP_ (Single nucleotide polymorphism-based heritability), IPC (Independent Pathway-Cholesky GRM-SEM model), ODD (Oppositional Defiant Disorder), r_g_ (genetic correlation).

### Sensitivity analysis

We carried out several sensitivity analyses. We (1) visually confirmed the similarity in structure between the best-fitting model and the bi-factor model across all analysed subsets (S_DLD_, S_LL_, S_ALL_, S_SSC_) (**Supplementary Table 4, Supplementary Figures 10-12**,**16**). Next, we (2) corroborated the superiority in model fit for all identified GRM-SEM models in SPARK and the SSC by comparing their fit with exploratory GRM-SEM models (**Supplementary Table 4**), such as one-factor independent pathway and IPC models (**Supplementary Figure 17**). To validate the predictive value of EFA models, we (3) confirmed the interchangeability of EFA methods predicting genetic factors (**Supplementary Tables 5-6**) and (4) found strong correlations between EFA-predicted and GRM-SEM estimated factor loadings (Pearson *r* > 0.98 for all analysed models, **Supplementary Figure 18**). Lastly, we (5) performed proof-of-principle simulations (**Supplementary Note 3**). We demonstrated the robustness of the proposed multi-step genomic covariance modelling approach (**Figure 1**), with little evidence for bias and sufficient 95% CI coverage for estimated factor loadings and derived variance components (**Supplementary Tables 11-14, Supplementary Figures 19-20**).

## DISCUSSION

Investigating genomic covariance across a broad spectrum of phenotypes in ASD using SEM-based techniques, this case-only study of two large autism cohorts demonstrates that the common genetic architecture of ASD is multi-dimensional. Here, we identified evidence for at least three independent common genetic dimensions associated with phenotypic heterogeneity in ASD.

For population-representative ASD, as reflected in SPARK, we identified three common genetic factors explaining predominantly variation in language/cognition, developmental delay and behavioural problems, with genetic dimensions essentially matching corresponding phenotypic measurements. For simplex ASD, within the SSC, we uncovered structural similarities supporting the first two factors (i.e. language/cognition and developmental delay), indicating conceptual replication. The major difference across cohorts concerned the genetic relationship between language/cognition and behavioural phenotypes. While genetic factors of language/cognition and behaviour were unrelated in population-representative ASD, the underlying phenotypes were strongly genetically related in simplex ASD and captured by a single dimension. Thus, profound structural differences exist in common genetic influences distinguishing population-representative and simplex ASD manifesting in ascertainment-specific patterns. Our findings strengthen the evidence for common genetic contributions to phenotypic variation in ASD (8,9,12) and offer insight into the underlying multi-dimensional common genetic architecture.

Across both cohorts, we found evidence for an independent language/cognition-related factor, as validated through association with higher liability to Asperger in SPARK. Although language performance is not included as a core symptom of ASD in the DSM-5 anymore, our findings confirm that autistic individuals differ considerably in their language presentation (30). While some children with ASD reach intact structural language skills, others are delayed or never master functional spoken language (30). Here, our analyses uncovered, through identification of the language factor, that genomic covariance between (higher) language level and (earlier) age of self-feeding with a spoon, an important personal-social developmental milestone which typically developing children master at about 15-18 months (31,32). Notably, the genetic influences contributing to the age by which children self-feed with a spoon were distinct from genetic factors underlying other motor developmental achievements, such as crawling, sitting or walking, when studied in SPARK. Infant autonomy in feeding, especially eating with the family, has been related to more advanced child language production and comprehension (33). Especially within SPARK (e.g. S_LL_ model), age of self-feeding with a spoon showed moderate to strong relationships with multiple language-related phenotypes and may present an early marker of cognitive and language development in ASD.

We also found robust evidence for a genetic factor that is related to developmental delay within SPARK and the SSC, explaining genetic variation underlying growth, such as the age of crawling, a developmental milestone children typically master between 9-18 months of age (34). Within SPARK, genetic variation was shared beyond the age of crawling (a proxy of the age of walking and sitting) across DCDQ motor control during movement (a proxy of DCDQ total score and fine motor handwriting), language disorder and RBSR self-injurious behaviour. These findings support the contribution of common genetic influences to variation in motor abilities, beyond association with *de novo* mutations (9), even if not captured by PGS for psychiatric disorders or PGS_EA_ (9). The spectrum of genetically linked developmental phenotypes, furthermore, extends reports of genetic associations between ASD polygenic risk and later age of walking in population-based samples (35).

Genetically mediated relationships between language/cognition phenotypes and behaviour across cohorts were heterogeneous, highlighting ascertainment-specific patterns. Within SPARK, the behavioural genetic dimension was independent of the language/cognitive dimension of influences. The behavioural-problems factor explained liability to ODD and variation in repetitive behaviour, especially RBSR sameness behaviour that is a proxy of RBSR total scores and ritualistic behaviour, but not self-injurious behaviour. We validated this factor through inverse genetic association with PGS_EA_, extending previous findings (9), independent of the language/cognitive dimension. In other words, educational attainment-related associations with symptom variation in ASD are unlikely to implicate cognitive factors, as captured by common genetic influences. Instead, our findings suggest that behavioural problems within a population-representative case-only ASD sample vary with non-cognitive correlates of socio-economic status. It is also possible that common genetic influences underlying the behavioural genetic dimension may, partially, tag rare variation given positive correlations between PGS_EA,_ and rare variant risk scores (9) in SPARK.

In contrast, within the SSC, we observed substantial genetic overlap between most language-related phenotypes and RBSR sameness behaviour. Simplex ASD, compared to multiplex ASD, is more often related to *de novo* mutations (11,14). Our findings may, therefore, present aetiological differences unique to simplex ASD, consistent with qualitative differences in the common genetic architecture of ASD individuals carrying *de novo* variants (5,6). Alternatively, genetic links between behaviour and language/cognition in the SSC might, to some degree, be a consequence of collider bias (36). Simplex families are recruited following strict ascertainment schemes (18). Collider bias can arise when two measures, such as behaviour and language/cognition, are independently related to a third variable, such as common genetic variation, and that third variable is conditioned upon (36). Here, the preferential ascertainment of simplex families depleted for inherited genetic risk (37), including common variation, may introduce artificial genetic relationships between behaviour and language/cognition. Stratifying SEM-predicted shared genetic factor structures by common, rare and *de novo* genetic architectures will shed further light on the complex links between genetic and phenotypic heterogeneity as part of future studies.

Our study has multiple strengths and limitations. First, we developed a data-driven GRM-SEM approach that utilises directly genotyped genome-wide information and facilitates building accurate multi-dimensional models of genomic covariance without the need for summary statistics. Here, we leverage genetic EFA to predict the genetic structure of the best-fitting GRM-SEM model, which is confirmed through comparison with a saturated model. Second, we demonstrate that the common genetic architecture of ASD is multi-dimensional. Thus, genetic analyses modelling the common genetic architecture of ASD require a sufficiently high number of phenotypes to allow for the empirical identification of these dimensions. Third, GRM-SEM relies on population-based assumptions of genotype distributions (i.e. Hardy-Weinberg equilibrium) and may exclude individuals or genetic variation that do not meet these expectations. Fourth, any genetic relationships within this study will reflect variation within an ASD case-only cohort. A mapping to external references, such as Asperger or PGS_EA_, can aid the interpretation of genetic factors across different research designs. Fifth, the lack of h^2^_SNP_ across phenotypes may not only reflect a lack of power but a lack of genetic heterogeneity across phenotypic variation in cases. Especially, social core phenotypes showed little evidence for h^2^_SNP_ possibly reflecting high social deficits across all studied individuals with ASD. Sixth, in this study we used transformed scores to aid model simplicity and the convergence of models. While we cannot exclude bias, given the robustness of sensitivity analyses and the consistency with previous findings, it is unlikely that transformed scores profoundly changed underlying genetic structures. Seventh, our study cannot yet address sex-specific differences in common genetic architectures, as previously reported (9), especially across non-European ancestry backgrounds. Because the prevalence of ASD is higher in males, the sex distribution in both samples is skewed. There is a low representation of females in ASD cohorts, given male preponderance of the condition, that prevents robust modelling using GRM-SEM and our results may, therefore, be less generalisable for females.

Together our results describe phenotypic variation in ASD as complex traits that are, at least partially, genetically linked due to common genetic factors that are augmented by ascertainment-specific patterns. Here, we show that multi-dimensional common genetic architectures can be accurately identified with a data-driven GRM-SEM approach utilising genome-wide genotyping data.

## ONLINE METHODS

### Samples

The SPARK cohort (https://sparkforautism.org/) (17) is a nationwide autism study across the US including simplex and multiplex families. Here, we studied SPARK phenotype (version 3) and genome-wide (version November 2018) data. This data freeze includes 59,218 individuals between ages 1 and 85, who received a professional diagnosis of ASD/autism (85%<18 years; 79% male), their biological parents, and, if available, one unaffected control sibling as well as all affected siblings for multiplex families (21,689 trios (including simplex families); 6,552 multiplex families). Written informed consent was completed by the parent or legal guardian of the children participating in the study.

The SSC cohort (https://www.sfari.org/resource/simons-simplex-collection/) (18) is a US collection of simplex families. Here, we investigated phenotype (version 15.3) and genome-wide (whole-genome 2 data release) data. This data freeze represents 2,591 affected children aged 4 to 17 years 11 months, including 2,643 simplex families with one (and only one) child with ASD and their unaffected biological parents and unaffected siblings. Informed consent and assent were provided for all participants.

We received ethical approval to access and analyse pre-collected de-identified genotype and phenotype data from these cohorts from the Radboud University Ethics Committee Social Science. All analyses were restricted to individuals with ASD with phenotypic and genetic information.

### Genotype information

#### SPARK

Genome-wide genotypes were obtained with the Infinium Global Screening Array-24 v.1.0. After individual and variant quality control (QC), 5,331 unrelated individuals (79.85% males, median age: 9 years) of European ancestry diagnosed with ASD, with genetic and phenotype information (see below) were included in the study (**Supplementary Methods 1, Supplementary Figure 1**). Individuals were excluded due to confirmed genetic syndromes/conditions, birth complications (i.e. birth defects, foetal alcohol syndrome, bleeding into the brain, insufficient oxygen at birth), other cognitive impairments or a brain injury (i.e. brain infection, lead poisoning, traumatic brain injury), similar to SSC exclusion criteria (see below). A genetic relationship matrix (GRM) (19) based on directly genotyped markers (N_SNPs_=450,491) was created in PLINK (v1.9) (38), applying a relationship cut-off of 0.05.

#### SSC

We used genome-wide data from three arrays: Illumina Human1M v1.0, Illumina Human1M-Duov3 and Illumina HumanOmni2.5. For each array, individual and variant QC were performed separately (see **Supplementary Methods 2**). Subsequently, genotype data were merged across the three arrays and again subjected to individual and variant-based QC (**Supplementary Methods 2**). After QC, 1,946 unrelated individuals (86.33% males, median age: 9 years) of European ancestry diagnosed with ASD with genetic and ASD phenotype information were included in the study (**Supplementary Figure 2**). Individuals were excluded according to SSC exclusion criteria, such as premature birth, brain injury/damage/abnormality, prenatal/birth complications, confirmed genetic syndromes/conditions, severe sensory/motor difficulties or nutritional/psychological deprivation. A GRM (19) based on directly genotyped markers (N_SNPs_=457,961) was created in PLINK (v1.9) (38), applying a relationship cut-off of 0.05.

### Phenotypes

#### SPARK

We studied parent-reported measures of ASD phenotypes and co-morbid disorders/disabilities spanning the domains of language and cognition (9 measures), general behaviour (9 measures), repetitive behaviour (7 measures), social (2 measures) and motor abilities (6 measures), as well as affective disorders (3 measures) and developmental milestones (11 measures). Phenotypes were extracted from the Basic Medical Screening Questionnaire (BMS), the Social Communication Questionnaire-Lifetime (SCQ) (39), the SPARK Background History Questionnaire (BGHX), the Repetitive Behaviours Scale-Revised (RBSR) (40), and the Developmental Coordination Disorder Questionnaire (DCDQ) (41), including 47 out of 149 available SPARK phenotypes (**Supplementary Methods 1, Supplementary Figure 1, Supplementary Table 1**).

The selected phenotypes included 21 categorical (within-sample prevalence of 5%) and 26 continuous phenotypes. At least 2,910 autistic individuals had phenotype and genotype data per trait (**Supplementary Table 1**). Among all the studied individuals in the SPARK sample, information on ASD subcategories was available for 1,754 individuals only: Asperger (N_ind_=716, 79.05% males, age range: 2-60 years), childhood autism (N_ind_=624, 81.57% males, age range: 1-55 years) and Pervasive Developmental Disorder Not Otherwise Specified (PDD-NOS, N_ind_=414, males=80.67% males, age range: 2-45 years). Consequently, we did not include ASD subcategory information directly within our modelling approach but instead mapped it onto the best-fitting model (reference Asperger=1, childhood autism=0, PDD-NOS=0, non-subcategory data= NA, deviance-transformed, see below).

#### SSC

We studied parent-reported measures of language and cognition (5 measures), general behaviour (1 measure), repetitive behaviour (4 measures), and motor abilities (3 measures), as well as developmental milestones (4 measures). These were comparable to SPARK measures, for follow-up analyses. Phenotypes were selected from the SSC BGHX, the SSC Diagnosis Summary Form, the SSC Medical History Interview, RBSR (40), DCDQ (41), the Child Behavior Checklist (CBCL 6-18) (42), and the Autism Diagnostic Observation Scale (ADOS) (43) (**Supplementary Figure 2, Supplementary Table 2**).

### Phenotype transformations

Continuous scores were transformed with ordinary least square regression and categorical scores with logistic regression [R:stats package]. Before transformation, all phenotypes were adjusted for sex, age, age squared, and ten ancestry-informative principal components (44), where the latter correct for subtle ancestry differences among individuals of Caucasian ancestry. For continuous phenotypes, residuals were rank-transformed and regressed again on covariates to achieve normality of transformed scores without a re-introduction of covariate effects (fully-adjusted two-stage rank normalisation)(45). For categorical phenotypes and co-morbid disorders, we constructed deviance residuals as the difference between the logistic model fit and the fit of an ideal model. Deviance residuals approximate the liability as observed within an ASD-only sample (henceforth referred to as liability), given that SPARK is a population-representative sample of ASD individuals, and there is no over-sampling of ASD cases with specific co-morbid disorders or phenotypes. For the SSC, liability is approximated for a simplex ASD case-only sample. We carried out extensive sensitivity analyses to ensure the validity of the transformed scores. For this purpose, we compared h^2^_SNP_ estimations (**Supplementary Figure 3**) and phenotypic correlation analyses for untransformed and transformed scores (**Supplementary Figure 21**). Pertinent to this work, analyses were conducted with transformed scores to ease the modelling process, i.e. rank-transformed scores for continuous phenotypes and deviance residuals for categorical phenotypes.

### Univariate and bivariate genetic variance analyses

Univariate (h^2^_SNP_) analyses, reflecting, here, the proportion of phenotypic variance among autistic individuals as explained by genotyped variants (SNPs), were carried out with GREML, as implemented in Genome-wide Complex Trait Analysis (GCTA, v1.93.0) software (19). GRMs were constructed from genome-wide genotyping information (see **Supplementary Methods 1-2**).

Bivariate genetic correlations (*r*_g_) across phenotypes were estimated using bivariate GREML (20). Genetic correlations reflect the extent to which the same genetic factors influence two measures.

### Multivariate modelling of genomic covariance

We modelled the multivariate genetic variance structure of ASD phenotypes using GRM-SEM as implemented in R:grmsem (v1.1.2, https://gitlab.gwdg.de/beate.stpourcain/grmsem) previously known as *gsem* (15,16).

GRM-SEM applies structural equation modelling techniques to analyse genomic covariance in samples of unrelated individuals using a maximum likelihood approach (15). We define the expected phenotypic variance, Σ_V_, of a multivariate normal phenotype Y (for 1…k traits) where Y_i_ ∼ N_k_ (μ, Σ_V_), as the sum of the expected genetic and residual variance components, Σ_A_ and Σ_E_:

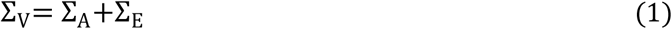

where Σ_V_, Σ_A_ and Σ_E_ are symmetric k x k matrices. The residual variance component, potentially, includes environmental factors, random error, non-additive genetic variance, rare variance or any other genetic influence not captured by the GRM (15,16,19). Within GRM-SEM, genetic and environmental influences are modelled as latent variables. The phenotypic variance for each measure Y can be dissected into genetic and residual influences (AE model), analogous to twin research (46):

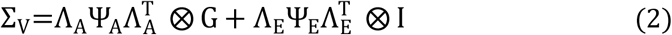

where Λ_A_ and Λ_E_ are matrices of genetic and residual factor loadings with dimensions k x p, where p is the number of factor loadings. Ψ_A_ and Ψ_E_ are p x p matrices of genetic and residual factor variances, respectively. G is a n x n GRM matrix for all pairs of n independent individuals constructed from the variants presented on a genome-wide genotyping chip, and I is a n x n identity matrix. The symbol ⊗ denotes the Kronecker product. In this work, Ψ_A_ and Ψ_E_ have been restricted to an identity matrix, given modest genetic correlations between latent variables, as predicted by oblique genetic EFA. Bi-factor models confirmed the independence of factor structures (see below). Note that we assume besides structured genetic covariance also structured residual covariance that can contribute to phenotypic covariance patterns (16). We analyse, here, a proportion of genetic variance in ASD individuals that can be modelled according to population-based principles.

We fitted the following multivariate models (15,16):

i. *Cholesky model*: The Cholesky decomposition model (**Supplementary Figure 17A**) is a saturated i.e. fully parametrised descriptive model without any restrictions on the structure of latent genetic and residual influences. This model is fitted to the data through the decomposition of both the genetic variance and residual variance into as many latent variables (factors) as there are observed variables. Here, Λ_A_ and Λ_E_ are k x k lower diagonal matrices. Note that other saturated models, such as direct symmetric models (47), were not fitted due to convergence problems with multicollinear data (not shown). Cholesky-derived genetic trait correlations provided input data to estimate the dimensionality of shared genetic factors (n_AC_) using genetic PCA (see below). The Cholesky-derived genetic trait covariance was used as input to predict the genomic covariance structure of the best-fitting model, with genetic EFA (see below).
ii. *Independent pathway model*: The independent pathway model (**Supplementary Figure 17B**) specifies one or more shared genetic and one or more shared residual factors, where n_AC_ is the number of shared genetic factors and n_EC_ is the number of residual factors, in addition to trait-specific genetic and residual influences, one for each trait. Λ_A_ and Λ_E_ have the dimensions k x p_a_ and k x p_e_, respectively, where p_a_ is the sum of n_AC_ and k, and p_e_ is the sum of n_EC_ + k. Pertinent to this study, we fitted 1-factor models only (n_AC_ = n_EC_ = 1).
iii. *Hybrid Independent Pathway/Cholesky model (IPC)*. The IPC model (**Supplementary Figure 17C**) structures the genetic variance as an independent pathway model (consisting of shared and measurement-specific influences where Λ_A_ has a dimension of k(n_AC_+k)) and the residual variance as a Cholesky model (where Λ_E_ is a lower diagonal k x k matrix). Here, we fitted 1-factor (n_AC_=1; k_traits_≥3) and multi-factor (n_AC_=2, k≥6; n_AC_=3; k≥8) IPC models, such that for the latter N_Λ(EFA)_<N_Λ(saturated)_. The genetic part of multi-factor IPC models was informed by the estimated number of genetic factors n_AC_ using proxy genetic PCA and the estimated factor loadings from genetic EFA (see below). Specifically, we used EFA-predicted information to define starting values and constraints (i.e. setting EFA factor loadings |λ|<0.1 in the corresponding genetic part of the GRM-SEM model to zero). As a rule of thumb, zero loadings have been defined as factor loading scores between -.10 and +.10 (48). Once fitted, we further trimmed the model by removing specific genetic factor loadings near zero (GRM-SEM factor loadings |λ|<0.01). The residual part of the model remained unchanged and was fitted as a Cholesky model.

To confirm the independence of shared genetic factors for the best-fitting multifactor IPC model, we fitted a bi-factor model of genetic variance within the IPC framework. The bi-factor model (49) consists of a general factor and one or more grouping factors, where each trait loads on the general factor, assuming statistical independence between these latent genetic dimensions. Given the bi-factor parametrisation, the model benefits from rotational invariance and unlimited dimensionality (50).

The goodness-of-fit for each model was evaluated with likelihood ratio tests (LRTs), Akaike information criterion (AIC) and the Bayesian information criterion (BIC) (51). Evidence for GRM-SEM factor loadings was assessed using Wald tests, based on unstandardized scores, while reported coefficients λ represent standardised factor loadings (setting the phenotypic variance to unit variance).

For the best-fitting models, we estimated heritability (h^2^_SNP_), genetic correlations (*r*_g_), and factorial co-heritabilities (f^2^_g_, i.e. the proportion of total trait genetic variance explained by a specific genetic factor). We defined bivariate genetic correlation between phenotypes, measuring the extent to which two phenotypes share genetic factors (ranging from -1 to 1 (52) according to:

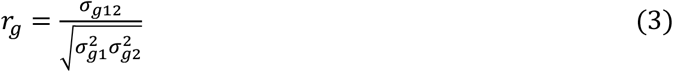

where σ_g12_ is the genetic covariance between two phenotypes 1 and 2, and σ^2^_g1_ and σ^2^_g2_ are the respective genetic variances. In addition, we estimate the factorial co-heritability f_g_^2^ as the relative contribution of a genetic factor to the genetic variance of a phenotype, defined as:

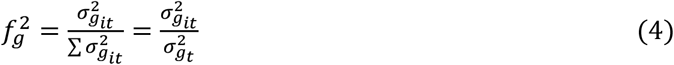

where σ^2^_g_it_ is the genetic variance of the genetic factor *i* contributing to trait t and σ^2^_g_t_ the total genetic variance of trait t, based on standardised factor loadings. Corresponding SEs were derived using the Delta method.

#### Phenotype selection

For GRM-SEM, we studied transformed phenotypes (see above) in combination with GRMs constructed from genotyped genome-wide variants in unrelated ASD individuals of European descent. GRM-SEM models are computationally expensive (15). For example, an 8-factor Cholesky decomposition model, as fitted within this study, can require up to 6 weeks of computing time even on a system incorporating at least four parallel cores of 3 GHz, and requiring up to 40 Gb (max vmem) memory. Hence, we streamlined the modelling process by combining measures of the same questionnaire (i.e. BGHX, DCDQ and RBSR) that shared an underlying genetic architecture (GRM-SEM *r*_g_=1). We only retained measures with the highest genetic correlations to (a) limit the number of studied phenotypes using proxy measures and (b) aid model convergence by reducing collinearity.

### Eigenvalue decomposition of genetic correlations: genetic PCA

The dimensionality of shared genetic factors (n_AC_) across a set of phenotypes was estimated by the spectral decomposition (53) [R:base package]. For the estimation, we used a Cholesky-derived genetic correlation matrix. Eigenvalues of this genetic PCA were plotted as a scree plot. The number of factors was estimated with the Optimal Coordinate criterion [R:nFactors package](54), applying a joint Kaiser’s rule (eigenvalue > 1) (55,56) and Cattell’s scree test (57).

### Exploratory factor analysis of genetic covariance: genetic EFA

Given evidence for multiple genetic factors (dimensionality of shared genetic factors n_AC_>1), we carried out for each set of selected phenotypes a genetic EFA (58) predicting underlying genetic factor structures, using *lavaan* (59) [R:lavaan package] software. As genetic trait covariance is not directly observable, we analysed the predicted genetic covariance matrix derived from a saturated (Cholesky) GRM-SEM model (see above). Factor solutions were estimated using a Diagonally Weighted Least Squares (DWLS) algorithm (60), i.e. a robust Weighted Least Squares (WLS) method that can be applied to skewed data where the likelihood function for any parameter θ is given as

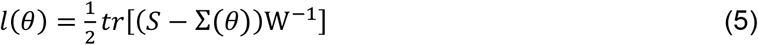

where S is the observed (here Cholesky predicted genetic covariance matrix) and Σ the EFA model-implied genetic covariance matrix. Inverse weighting was carried out with a diagonal weight matrix W, based on the estimated variance 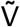 of the genetic covariance V_A_, as derived with a Cholesky model, where 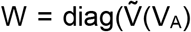. For comparison, we also carried out an unweighted least square estimation, where the identity matrix replaces W. Factors in *lavaan* were rotated using either orthogonal or oblique rotation techniques, performing EFA varimax and oblimin, respectively. We opted for an EFA varimax model if the predicted genetic correlation between genetic factors by an EFA oblimin model was modest (i.e. *r*≤0.32 (21) and thus ignorable) or if the EFA oblimin model produced a similar pattern of loadings as EFA varimax (21). In other words, the EFA oblimin solution did not increase the simplicity of the model (21). The factor loadings of the selected EFA model were utilised to define starting values and constraints of GRM-SEM multifactor IPC models, setting genetic EFA factor loadings |λ|<0.1 in the corresponding genetic part of the GRM-SEM model to zero (21).

Note that an evaluation of EFA models based on model fit criteria established in observational research is not meaningful here, as the studied genetic covariance matrix (Cholesky) is estimated with an error that may result in negative uniqueness of the predicted genetic variance, violating modelling assumptions (known as a Heywood case) (61).

For sensitivity analyses, we also compared estimates of EFA *lavaan* with estimates of other EFA software such as *fa* [R:psych package] which does not allow for inverse weighting (62).

### Simulation study

To evaluate the robustness of the proposed multi-step genomic covariance modelling approach, and in particular to assess bias, we carried out simulations comparing true values with GRM-SEM IPC factor loadings, but also EFA-predicted factor loadings (**Supplementary Tables 11-14, Supplementary Figures 19-20**), as described in detail in the supplement (**Supplementary Note 3**).

In brief, assuming multivariate normality, we simulated six-variate traits with either two shared genetic factors without correlation or two shared genetic factors with cross-loading as detailed by path models in **Supplementary Figures 19 and 20**, respectively, across 20 replicates. Each six-variate trait was based on Z-standardized phenotypes with 2,000 individuals per phenotype and (for simplicity) 5,000 causal loci, to increase power. Besides the median estimate, simulation performance measures included the median bias, the median empirical standard error (empSE) and coverage of 95%-confidence intervals (such that the estimated 95%-confidence interval contains the true value), and the respective Monte-Carlo SEs (MCSE).

### Multiple testing

A correction for multiple testing of estimated GRM-SEM factor loadings of our analysis is not directly applicable. We jointly analyse multiple phenotypes using a multivariate approach to comprehensively represent all shared genetic factors across the studied phenotypic spectrum. h^2^_SNP_ and r_g_ estimates from a GCTA screen within Stage I are not individually interpreted, given the preliminary character of these analyses. However, if a multiple testing adjustment for individual measures reported during Stage I were considered, an experiment-wide threshold of *p*<0.0015 (0.05/34 independent measures) would be needed to be applied, as estimated with Matrix Spectral Decomposition (matSpD) (63), based on phenotypic score correlations.

### Univariate polygenic scoring analysis in SPARK

Consistent with current guidelines (64), we constructed PGS for EA within SPARK based on high-quality genome-wide imputed SNPs (**Supplementary Methods 3**), utilising available summary statistics from recent EA meta-GWAS (65). For this purpose, we used PRS-CS software (66), which applies continuous-shrinkage parameter to adjust SNP effect sizes for linkage disequilibrium. Once SNP effect sizes were calculated in PRS-CS, PGS_EA_ scores were calculated in PLINK (38) and, subsequently, Z-standardised.

## Data Availability

Genotype and phenotype data from the SPARK and SSC cohorts are available upon application and approval from the Simons Foundation Autism Research Initiative (SFARI) (https://www.sfari.org/resource/autism-cohorts/). Approved researchers can obtain the SPARK and SSC population dataset described in this study by applying at https://base.sfari.org. GWAS summary statistics for educational attainment were accessed through the Social Science Genetic Association Consortium (SSGAC, Login via https://thessgac.com/).

## DATA AVAILABILITY

Genotype and phenotype data from the SPARK and SSC cohorts are available upon application and approval from the Simons Foundation Autism Research Initiative (SFARI) (https://www.sfari.org/resource/autism-cohorts/). Approved researchers can obtain the SPARK and SSC population dataset described in this study by applying at https://base.sfari.org. GWAS summary statistics for educational attainment were accessed through the Social Science Genetic Association Consortium (SSGAC, https://thessgac.com/).

## CODE AVAILABILITY

In this study, we used the following software packages: PLINK (PLINK v1.9, https://www.cog-genomics.org/plink/1.9/), PRScs (https://github.com/getian107/PRScs), GCTA-GREML (GCTA v1.93, https://cnsgenomics.com/). We used the following R packages: stats 4.0.2, base 4.0.2, nFactors 2.4.1, psych 2.2.3, lavaan 0.6-10, grmsem 1.1.2 (https://gitlab.gwdg.de/beate.stpourcain/grmsem).

## ACKNOWLEDGMENTS

We are grateful to all of the families in SPARK, the SPARK clinical sites and SPARK staff. We are grateful to all of the families at the participating Simons Simplex Collection (SSC) sites, as well as the principal investigators (A. Beaudet, R. Bernier, J. Constantino, E. Cook, E. Fombonne, D. Geschwind, R. Goin-Kochel, E. Hanson, D. Grice, A. Klin, D. Ledbetter, C. Lord, C. Martin, D. Martin, R. Maxim, J. Miles, O. Ousley, K. Pelphrey, B. Peterson, J. Piggot, C. Saulnier, M. State, W. Stone, J. Sutcliffe, C. Walsh, Z. Warren, E. Wijsman). We appreciate obtaining access to phenotypic and genetic data on SFARI Base. This work was supported by a grant from the Simons Foundation Autism Research Initiative (SFARI ID: 514787, PI B.S.P.) covering the work of L.H., M.B. and M.M.J.D, in addition to support from the Max Planck Society. F.S., E.V., S.E.F. and B.S.P. were fully supported by the Max Planck Society. C.Y.S. was supported by the UK Medical Research Council (MRC) Integrative Epidemiology Unit at the University of Bristol (MC_UU_00011/3). J.B. was supported by the EU-AIMS and AIMS-2-TRIALS programmes which receive support from Innovative Medicines Initiative Joint Undertaking Grant No. 115300 and 777394, the resources of which are composed of financial contributions from the European Union’s FP7 and Horizon2020 Programmes, and from the European Federation of Pharmaceutical Industries and Associations (EFPIA) companies’ in-kind contributions, and AUTISM SPEAKS, Autistica and SFARI; and by the Horizon2020 supported programme CANDY (Grant No. 847818).

