## Supplementary Material for "Structural models of genome-wide covariance identify multiple common dimensions in autism"

### SUPPLEMENTARY NOTES

#### SUPPLEMENTARY NOTE 1. PHENOTYPIC SUBSET IDENTIFICATION AND GENETIC PROXY MEASURES

To facilitate model convergence, we retained GCTA genetic correlations across ASD phenotypes that passed the p-value threshold of $p(r_g) \leq 0.1$ (**Figure 2B**, **Supplementary Figure 4**). Based on the retained genetic correlations patterns, we selected subsets of phenotypes in SPARK for in-depth multivariate modelling (**Supplementary Table 3**).

Sequentially, we studied the genetic correlations of each of the 17 phenotypes with the other phenotypes. We observed four phenotype subsets implicating language/cognition-related phenotypes (**Supplementary Table 3**). These included two large phenotype subsets related to (1) language disorder and (2) language level, as well as two smaller subsets related to (3) cognitive age level and (4) language age level. Regarding behavioural problems and repetitive behaviour, we identified three phenotype subsets involving (5) oppositional defiant disorder (ODD), (6) Repetitive Behaviours Scale-Revised (RBSR) total score and (7) RBSR self-injurious scores. There was a single motor subset capturing genetic links with Developmental Coordination Disorder Questionnaire (DCDQ) proxies (8) as well as two phenotype subsets linked to developmental milestones, implicating the age of self feeding (9) and the age of crawling (10).

To ensure the stability of the genetic models and to avoid collinearity problems, once the phenotype subsets were identified, highly correlated phenotypes measured with the same questionnaire were reduced, retaining only a single representative measure, or proxy. To select proxy measures, we first fitted one-factorial GRM-SEM IPC models (**Supplementary Figure 5**) across scales within the same questionnaire to identify genetically similar measures (GRM-SEM $r_g$=1). Across developmental milestones (SPARK Background History Questionnaire, BGHX, **Supplementary Figure 5A-C**), except for the age at which children started to self-feed with a spoon (age of self feeding) scores genetically proxied each other.

Across motor phenotypes (DCDQ, **Supplementary Figure 5D-F**), genetic variation was shared and thus, scores genetically proxied each other. These findings were similar for repetitive behaviour scales (RBSR, **Supplementary Figure 5G-I**), except for a small specific genetic variance contribution to self-injurious behaviour.

We started the model-building process by selecting three phenotype subsets related to developmental language disorder/delay (here forth referred to as language disorder, $S_{DLD}$), language level ($S_{LL}$) and age of crawling ($S_{CRL}$), respectively, that comprehensively represent all other combinations of related measures (**Figure 2B**, **Supplementary Table 3**).

#### SUPPLEMENTARY NOTE 2. STRUCTURAL MODELS OF $S_{DLD}$, $S_{LL}$, $S_{ALL}$ AND $S_{SSC}$

To model the multivariate genetic architecture as captured by the selected phenotype subsets or combinations thereof, we carried out a series of genetic principal component analysis (PCA) eigenvalue decompositions, genetic exploratory factor analysis (EFA) steps and, finally, confirmatory factor analysis (GRM-SEM) steps (**Figure 1**, **Methods**). Across all subsets, EFA oblimin predicted factor correlations were modest (**Supplementary Table 5**), and GRM-SEM models were, thus, informed by EFA varimax predictions (**Supplementary Table 6**). Each of the presented models below fitted the data best, based on AIC and BIC, with a fit that was comparable to a saturated (Cholesky) model and a bi-factor model (**Table 1**, **Supplementary Table 4**), where the latter confirmed the predicted independence of genetic factors.

For the $S_{DLD}$ phenotype subset (**Figure 3A-C**, **Supplementary Table 7**, **Supplementary Figure 6**), we investigated genomic covariance across three language/cognition phenotypes (language disorder, cognitive age level, language level), general behaviour (ODD), a developmental milestone (age of self feeding), a motor score proxy (DCDQ total score) and a repetitive behaviour proxy (RBSR total score). Eigenvalue decomposition of Cholesky-derived genetic trait correlations identified two genetic dimensions (**Figure 3A**) that were modestly genetically correlated ($r$=0.26), consistent with factorial independence. The best-fitting model, based on AIC and BIC, was an IPC model with two genetic factors (**Table 1**), closely matching the fit of the saturated model ($p_{LRT}$=1). We identified a common genetic factor accounting, predominantly, for variation in language/cognition phenotypes ($A_{lang}$) with the strongest factor loadings for language level ($\lambda_{lang}$=0.47,SE=0.08), and a factor explaining variation in behavioural-problems ($A_{beh}$) with the strongest factor loading for liability to ODD ($\lambda_{beh}$=0.41,SE=0.09) (**Figure 3B-C**).

The $S_{LL}$ phenotype subset (**Figure 3D-F**, **Supplementary Table 8**, **Supplementary Figure 7**) comprised four language/cognition phenotypes (language disorder, cognitive age level, language age level, language level), two developmental milestones (age of crawling, age of self feeding) and the self-injurious repetitive behaviour sub-phenotype (RBSR self-injurious). Genetic PCA eigenvalue decomposition of Cholesky-derived genetic trait correlations identified two genetic dimensions (**Figure 3D**) with little correlation (*r*=-0.03). Fitting structural models, informed by genetic EFA, revealed that a two-factor IPC model fitted the data best (**Table 1**), based on AIC and BIC, with a fit close to the saturated model ($p_{LRT}$=1). The first genetic factor explained, predominantly, language/cognition phenotypes ($A_{lang}$), with the strongest factor loadings for language level ($\lambda_{lang}$ =0.44,SE=0.08), as observed for $S_{DLD}$. However, the second genetic factor accounted for variation in developmental-delay ($A_{dev}$), not behavioural-problems, with the strongest factor loading for age of crawling ($\lambda_{dev}$=0.46,SE=0.12, **Figure 3E-F**).

For the third phenotype subset, age of crawling ($S_{CRL}$), genetic PCA eigenvalue decomposition did not reveal a clear factor dimension (**Supplementary Figure 8**). Therefore, in line with our study design, we proceeded to combine phenotypes from the three subsets ($S_{DLD}$,$S_{LL}$,$S_{CRL}$) into a joint set ($S_{ALL}$), facilitating the identification of a larger overarching genetic structure. We retained phenotypes with the strongest factor loadings as well as cross-loadings from $S_{DLD}$ and $S_{LL}$ and combined them with all $S_{CRL}$ phenotypes (**Supplementary Table 3**). Due to computational limitations, we excluded two $S_{LL}$ phenotypes (cognitive age level and language age level) that were solely related to the language/cognition factor (**Supplementary Figure 6**, **Supplementary Figure 7**) factor and highly correlated with the remaining measures (cognitive age level, language level: GCTA $r_g$=0.87,SE=0.29; language age level, language level: GCTA $r_g$=1.00,SE=0.24, **Figure 2B**), showing similar association patterns in structural models (**Supplementary Figure 6, 7**).

The $S_{ALL}$ phenotype subset (**Figure 3G-I**, **Supplementary Table 9**, **Supplementary Figure 9**) comprised two language/cognition phenotypes (language disorder, language level), general behaviour (ODD), two developmental milestones (age of crawling, age of self feeding), a DCDQ motor proxy (control during movement) and two RBSR repetitive behaviour scores (self-injurious behaviour, sameness behaviour). Genetic PCA eigenvalue decomposition of Cholesky-derived genetic trait correlations identified three genetic dimensions (**Figure 3G**) with modest correlation (*r*=-0.34-0.06). Fitting structural models, informed by genetic EFA, revealed that a three-factor IPC model fitted the data best (**Table 1**), based on AIC and BIC, with a fit close to the saturated model ($p_{LRT}$=1). The first genetic factor explained, predominantly, language/cognition phenotypes ($A_{lang}$), with the strongest factor loadings for language level ($\lambda_{lang}$=0.46,SE=0.08), as observed for $S_{DLD}$ and $S_{LL}$. The second genetic factor accounted for variation in developmental-delay ($A_{dev}$), with the strongest factor loading for age of crawling ($\lambda_{dev}$=0.47,SE=0.10), as observed for $S_{LL}$. The third genetic factor accounted for variation in behavioural-problems ($A_{beh}$), with the strongest factor loading for liability to ODD ($\lambda_{beh}$=0.45,SE=0.09), as observed for $S_{DLD}$.

The $S_{SSC}$ phenotype subset (**Figure 6**, **Supplementary Table 10**) comprised three language/cognition phenotypes (language disorder, language age level, language level), general behaviour (ODD), three developmental milestones (age of crawling, age of self feeding, age of walking), and the RBSR repetitive behaviour score (sameness behaviour). Eigenvalue decomposition of Cholesky-derived genetic trait correlations identified three genetic dimensions (**Figure 6A**) with small correlation (*r*=-0.12-0.13). Fitting structural models, informed by genetic EFA, revealed that a three-factor IPC model fitted the data best (**Table 1**), based on AIC and BIC, with a fit close to the saturated model ($p_{LRT}$=1). The first genetic factor ($A_{F1}$) explained, predominantly, language/cognition phenotypes, with the strongest factor loadings for age of self feeding ($\lambda_{F1}$=-0.46,SE=0.19), as observed for the $A_{lang}$ factor in $S_{ALL}$. The second genetic factor ($A_{F2}$) accounted for variation in developmental-delay, with the strongest factor loading for age of walking ($\lambda_{F2}$=0.62,SE=0.14), as observed for the $A_{dev}$ factor $S_{ALL}$. The third genetic factor ($A_{F3}$) accounted for variation that is shared across language/cognition, but also repetitive (RBSR sameness) behaviour, with the strongest factor loading for language age level ($\lambda_{F3}$=0.61,SE=0.10).

#### SUPPLEMENTARY NOTE 3. GRM-SEM MULTI-FACTOR MULTIVARIATE MODELLING APPROACH AND SIMULATIONS

Data-driven genomic covariance modelling using GRM-SEM consisted of a multiple-step approach, as outlined in **Figure 1**, with each step being described in full within the Methods section: We fitted (i) a saturated GRM-SEM (Cholesky) model to describe the genetic architecture in full. Based on this information, we (ii) predicted the number of shared genetic factors ($n_{AC}$) across phenotypes through eigenvalue decomposition of Cholesky-derived genetic correlations. If $n_{AC}$>1, we (iii) approximated the underlying genetic factor structure through exploratory factor analysis (EFA) of Cholesky-derived genetic trait covariance. We used the information from (ii) and (iii) to fit (iv) a series of multi-factor Independent Pathway/Cholesky (IPC) models, including bi-factor models (to confirm the independence of shared genetic factors). For comparison, we also fitted one-factor Independent Pathway (IP) and IPC models (v). We compared (vi) the model fit of multi-factor models to one-factor models and the saturated (Cholesky) model to identify the best-fitting model.

To confirm the robustness of the proposed data-driven genomic covariance modelling approach, we conducted simulations from a parametric model assessing evidence for bias. Path diagrams depicting a multi-factorial six-variate trait consisting of two shared genetic factors without (scenario 1) and with (scenario 2) cross-loadings are shown in **Supplementary Figures 19A and 20A**, respectively. The true values for path coefficients and corresponding genetic and residual variances are given in **Supplementary Tables 11-14**. To reduce the computational burden, we assumed 2,000 individuals per trait and 5,000 causal loci conducting 20 simulations per scenario (**Supplementary Figures 19A and 20A**).

Besides the median estimate ($\hat{\beta}$) representing either path coefficients ($\hat{\lambda}$) or derived variances ($\hat{\theta}$), simulation performance measures were median bias, median empirical standard error (empSE) and coverage of 95%-confidence intervals (such that the estimated 95%-confidence interval contains the true value $\beta$) and the respective Monte-Carlo SEs (MCSE), as defined below (1):

$$bias = \frac{1}{N_{sim}}\sum_{i=1}^{N_{sim}}(\widehat{\beta_i} - \beta), \qquad MCSE\ bias = \sqrt{\frac{1}{(N_{sim}-1)N_{sim}}\sum_{i=1}^{N_{sim}}\left(\widehat{\beta_i} - \bar{\beta}\right)^2} \tag{1}$$

$$EmpSE = \sqrt{\frac{1}{(N_{sim}-1)}\sum_{i=1}^{N_{sim}}\left(\widehat{\beta_i} - \bar{\beta}\right)^2}, \qquad MCSE\ EmpSE = \frac{\widehat{EmpSE}}{\sqrt{2(N_{sim}-1)}} \tag{2}$$

$$Coverage = \frac{1}{N_{sim}}\sum_{i=1}^{N_{sim}} 1(\hat{\beta}_{l,i} \leq \beta \leq \hat{\beta}_{u,i}), \qquad MCSE\ Coverage = \sqrt{\frac{\widehat{Coverage}(1-\widehat{Coverage})}{N_{sim}}} \tag{3}$$

Simulations predicted two underlying genetic factors from Cholesky-derived genetic trait correlations, throughout. IPC starting values and constraints for the genetic part were obtained from two-factor EFA varimax *lavaan* models fitted to a Cholesky-estimated genetic variance/covariance matrix (with diagonal inverse variance weights based on standard errors), given that EFA oblimin predicted correlations between genetic factors were near zero (scenario 1: median EFA-predicted correlation: -0.020; scenario 2: median EFA-predicted correlation: $<-10^{-10}$). Subsequent GRM-SEM models were, thus, exclusively informed by EFA varimax factor predictions. EFA-derived genetic factor loadings <0.1 were constrained to zero. Among the 18 possible genetic path coefficients (based on a 2-factor IPC model), the multi-step approach accurately identified zero and non-zero parameters with bias ranging between -0.035 to 0.007 for a six-variate trait without cross-loading (scenario 1, **Supplementary Table 11**), and with bias between -0.030 to 0.015 for a six-variate trait with cross-loading (scenario 2, **Supplementary Table 13**). Consistent with the simulated trait architecture, on average, 33 genetic and residual path coefficients were estimated for scenario 1 (without cross-loading, **Supplementary Table 11**, median bias range: -0.035 to 0.022) and 34 path coefficients for scenario 2 (with cross-loading, **Supplementary Table 13**, median bias range: -0.030 to 0.030), confirming the robustness of the method. Coverage of path coefficients was sufficient and, taking MCSE into account, consistent with 95% or higher probability that confidence intervals contain the true value across estimated parameters, except one parameter in scenario 1 (94% probability). For derived genetic and residual trait covariance, based on the modelled six-variate traits in scenario1 and 2, median bias and coverage of true values were similar (**Supplementarys Table 12 and 14**).

### SUPPLEMENTARY METHODS

#### SUPPLEMENTARY METHODS 1. GENOTYPE QUALITY CONTROL IN THE SPARK COHORT

Genetic data for the SPARK cohort were based on the SPARK November 2018 release (2) (Infinium Global Screening Array-24 v.1.0; $N_{SNPs}$=632,015; $N_{ind}$=27,064). Genotypes were lifted from Build38 to Build37 (hg19) and subjected to quality control (QC).

As part of individual QC measures, we excluded individuals due to sex mismatch, duplicated individuals, individual missingness (>3% missing data), non-European ancestry (MDS analysis; utilising all 1,000 Genomes Phase 3 populations as reference for clustering) and SPARK exclusion criteria. In line with SSC exclusion criteria, we excluded SPARK individuals with reported genetic diagnoses from the SPARK Basic Medical Measures questionnaire (e.g. Rett's Syndrome, DiGeorge syndrome, Fragile X) or due to serious environmental complications affecting central nervous system development (spina bifida, foetal alcohol syndrome, insufficient oxygen at birth, bleeding into the brain, serious prenatal infection, brain infection, lead poisoning or traumatic brain injury) or cognitive delays or impairment due to another medical condition or exposure (e.g. brain injury, stroke, lead poisoning, HIV, radiation, hydrocephalus, brain tumour, drug effects). The sample was eventually restricted to ASD probands only.

Variant QC excluded SNPs with a SNP missingness >5%, violations of Hardy-Weinberg equilibrium ($p$<5x10$^{-7}$), minor allele frequency (MAF) < 1% as well as non-autosomal SNPs.

After QC, a genetic relationship matrix (GRM) of 450,491 autosomal variants was created in PLINK applying a relationship cut-off of 0.05, based on a total of 5,331 individuals.

#### SUPPLEMENTARY METHODS 2. GENOTYPE QUALITY CONTROL IN THE SSC COHORT

Genetic data for the SSC cohort were based on the SSC Whole-genome 2 data release, including data from 3 arrays: Illumina Human1M v1.0 ($N_{SNPs}$=1,072,841; $N_{ind}$=1,354), Illumina Human1M-Duov3 ($N_{SNPs}$=1,199,033; $N_{ind}$=4,626), Illumina HumanOmni2.5 ($N_{SNPs}$=2,440,283; $N_{ind}$=4,240). Genotypes were lifted from Build 36 to Build 37 (hg19) and subjected to quality control (QC). For each array, individual and variant QC was carried out separately.

As part of individual QC measures, we excluded individuals due to sex mismatch, individual missingness (>3% missing data), non-European ancestry (MDS analysis; utilising all 1,000 Genomes Phase 3 populations as reference for clustering) and SSC exclusion criteria. The sample was eventually restricted to ASD probands only.

Variant QC excluded SNPs with a SNP missingness > 5% and violations of Hardy-Weinberg equilibrium ($p<5x10^{-7}$). Subsequently, genotype data were merged across the three arrays ($N_{SNPs}$= 2,757,032; $N_{ind}$= 1,966) and subjected to additional QC measures.

Additional QC measures on the merged file excluded individuals with >3% missing data and non-European ancestry (as described above). Furthermore, SNPs with >5% missingness, violations of HWE ($p<5x10^{-7}$), MAF<1% and non-autosomal SNPs were excluded.

After QC, a genetic relationship matrix (GRM) of 457,961 autosomal variants was created in PLINK applying a relationship cut-off of 0.05, based on a total of 1,946 individuals.

#### SUPPLEMENTARY METHODS 3. UNIVARIATE POLYGENIC SCORING ANALYSIS IN THE SPARK COHORT

To enhance the interpretability of identified genetic structures, we mapped ASD subcategory information and polygenic scores (PGS) for educational attainment (EA) onto the model structure of the $S_{ALL}$ model in SPARK. To compute $PGS_{EA}$ we used PRS-CS (3), a Bayesian-based approach that adjusts SNP effect sizes for linkage disequilibrium (LD) by applying a continuous-shrinkage parameter. The auto-option for a fully Bayesian estimation of the shrinkage parameter phi was selected. Furthermore, we used the software's default settings: 'a' in the gamma-gamma prior to 1, 'b' in the gamma-gamma prior to 0.5 and selecting 1,000 Markov Chain Monte Carlo iterations, 500 burn-in iterations, and a Markov chain thinning factor of 5. As LD reference file, we used the UK Biobank European reference panel recommended on the software's git-hub page (https://github.com/getian107/PRScs).

Summary statistics for educational attainment (4) were obtained from the SSGAC repository (https://thessgac.com/). The log odds of genetic SNP effects were aligned to indicate alleles with increased educational attainment. Once SNP effect sizes were calculated in PRS-CS, $PGS_{EA}$ scores were calculated in PLINK(5) and, subsequently, Z-standardised.

$PGS_{EA}$ were constructed for 5,331 unrelated SPARK individuals (genomic relatedness<0.05, see **Supplementary Methods 1** for more details), based on high-quality imputed SNPs (INFO>0.8, 95%-posterior genotyping probability>0.9, MAF>0.005).

### SUPPLEMENTARY FIGURES

**Supplementary Figure 1.** Individual and phenotype selection in the Simons Foundation Powering Autism Research for Knowledge (SPARK) sample. 14
**Supplementary Figure 2.** Individual and phenotype selection in the Simons Simplex Collection (SSC) sample. 15
**Supplementary Figure 3.** GCTA $h^2_{SNP}$ estimates of screened SPARK phenotypes ($N_{pheno}$=47). 16
**Supplementary Figure 4.** Genetic correlations ($r_g$) across phenotypes in SPARK. 17
**Supplementary Figure 5.** Proxy measure identification in SPARK. 18
**Supplementary Figure 6.** Best-fitting two-factor GRM-SEM model for the language disorder ($S_{DLD}$) set in SPARK. 19
**Supplementary Figure 7.** Best-fitting two-factor GRM-SEM model for the language level ($S_{LL}$) set in SPARK... 20
**Supplementary Figure 8.** Scree plot for the age of crawling ($S_{CRL}$) set in SPARK. 21
**Supplementary Figure 9.** Best-fitting three-factor GRM-SEM model for the combined ($S_{ALL}$) 22
**Supplementary Figure 10.** Bi-factor GRM-SEM model for the language disorder ($S_{DLD}$) set in SPARK. 23
**Supplementary Figure 11**. Bi-factor GRM-SEM model for the language level ($S_{LL}$) set in SPARK. 24
**Supplementary Figure 12.** Bi-factor GRM-SEM model for the combined ($S_{ALL}$) set in SPARK. 25
**Supplementary Figure 13.** Heritability comparison between GCTA and GRM-SEM. 26
**Supplementary Figure 14.** GCTA $h^2_{SNP}$ estimates of dichotomised ASD subcategories in SPARK. 27
**Supplementary Figure 15**. GCTA $h^2_{SNP}$ estimates of Simons Simplex Collection (SSC) phenotypes (follow-up). 28
**Supplementary Figure 16.** Follow-up bi-factor GRM-SEM model in the SSC ($S_{SSC}$). 29
**Supplementary Figure 17.** GRM-SEM multivariate model types. 30
**Supplementary Figure 18.** Comparison of factor loadings estimated with EFA lavaan and GRM-SEM. 31
**Supplementary Figure 19.** GRM-SEM simulations of a six-variate trait with two independent common genetic factors without cross-loadings. 32
**Supplementary Figure 20.** GRM-SEM simulations of a six-variate trait with two independent genetic factors with cross-loading. 33
**Supplementary Figure 21.** Phenotypic correlations of ASD phenotypes in SPARK. 34

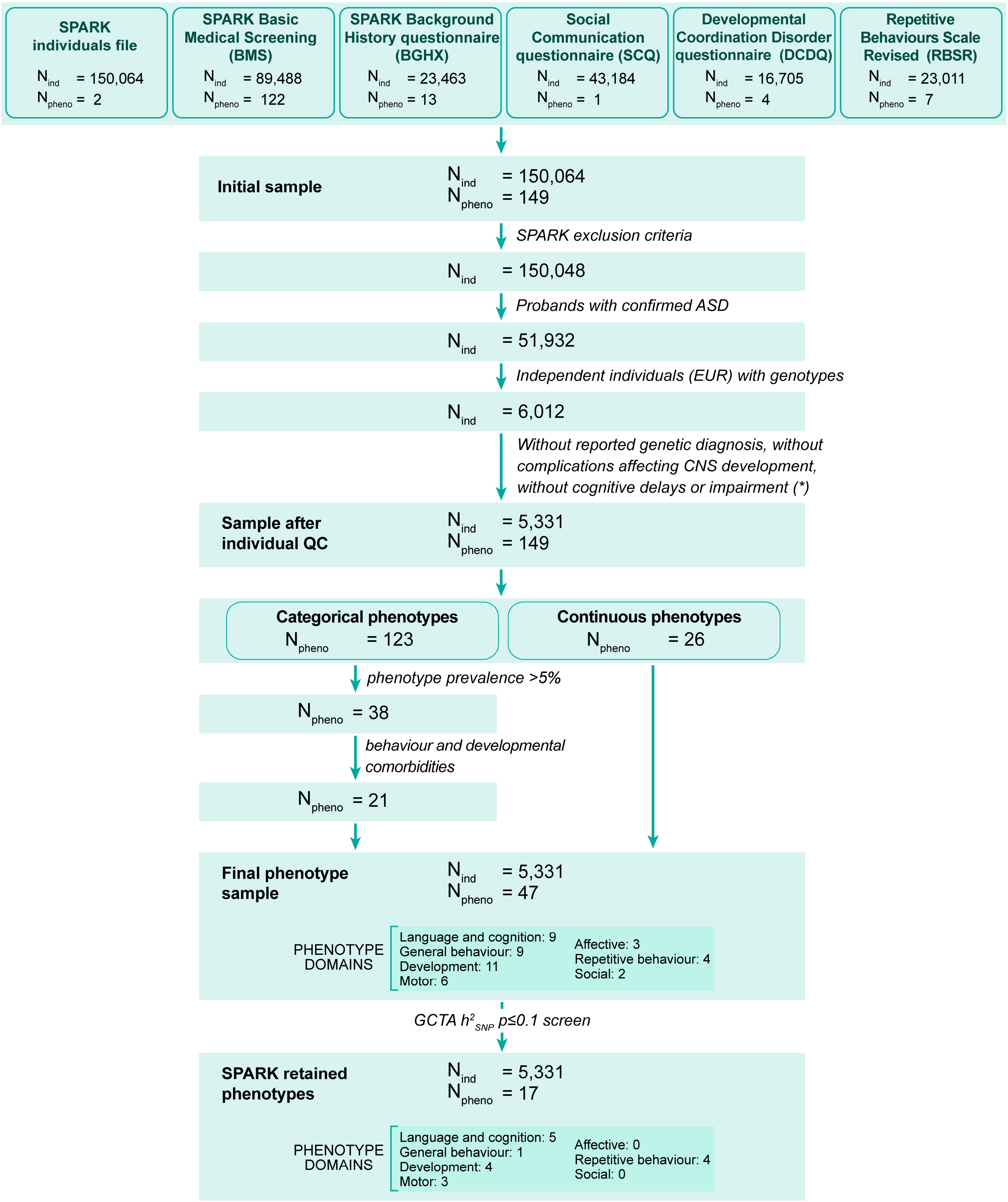

**Supplementary Figure 1. Individual and phenotype selection in the Simons Foundation Powering Autism Research for Knowledge (SPARK) sample.** (*) See Supplementary Methods 1 for a more detailed description.

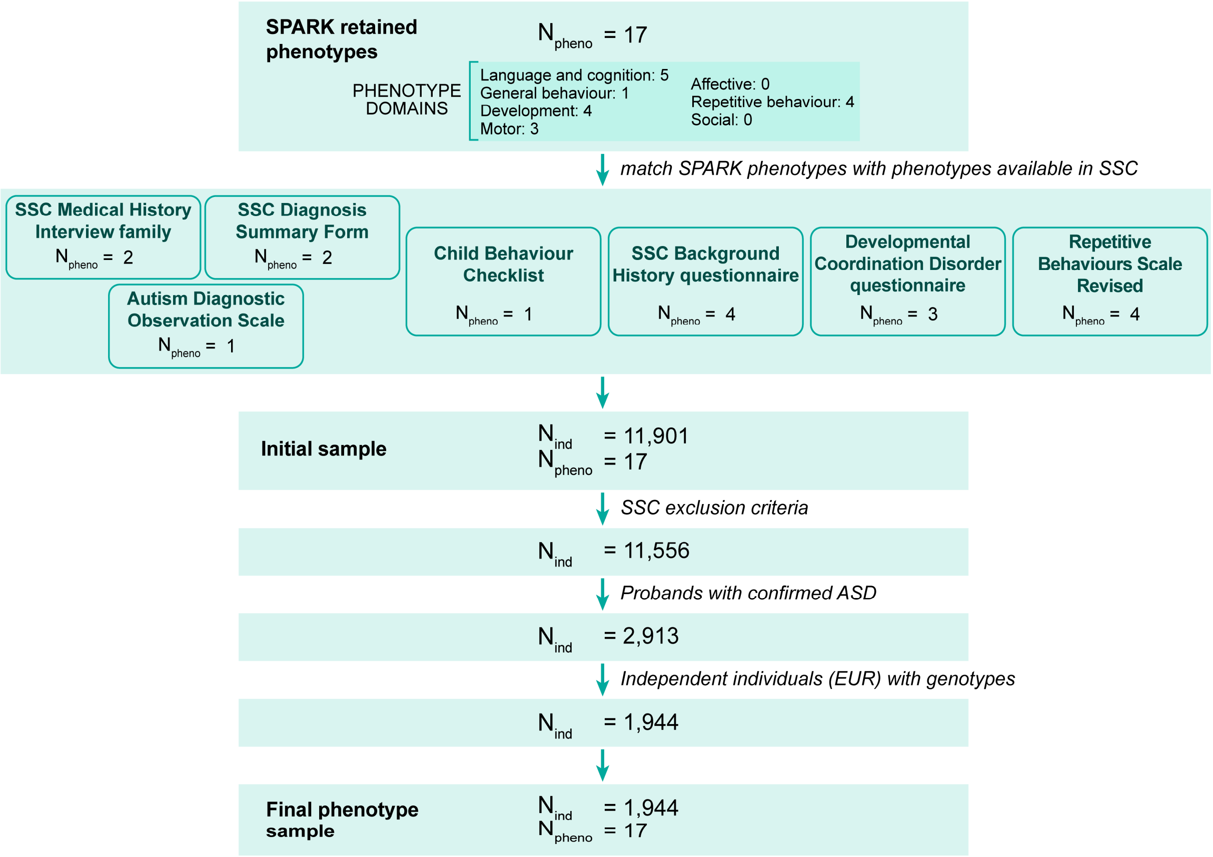

**Supplementary Figure 2. Individual and phenotype selection in the Simons Simplex Collection (SSC) sample.** Individual and phenotype selection in the SSC sample. 17 phenotypes were selected, analogous to 17 phenotypes retained in SPARK (Figure 2), from the SSC Medical History Interview (MEDHX fam, 2 language/cognition phenotypes), the SSC Diagnosis Summary Form (2 language/cognition phenotypes), Autism Diagnostic Interview-Revised (ADI-R, a language/cognition phenotype), the Child Behaviour Checklist (CBCL 6-18, a behavioural phenotype), the SSC Background History Questionnaire (BGHX, 4 developmental phenotypes), the Developmental Coordination Disorder Questionnaire (DCDQ, 3 motor phenotypes), and the Repetitive Behaviours Scale-Revised (RBSR, 4 repetitive behaviour phenotypes).

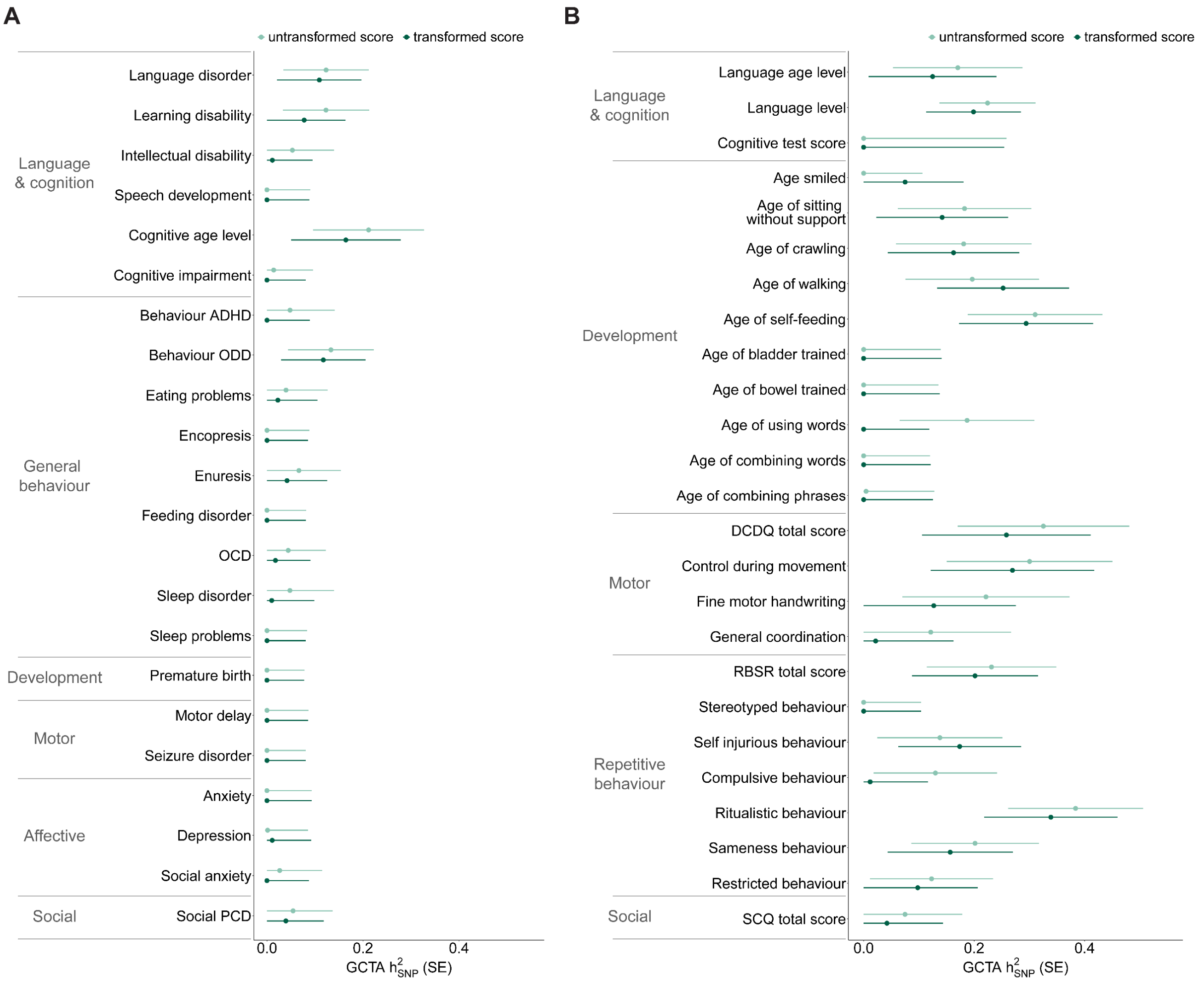

**Supplementary Figure 3. GCTA $h^2_{SNP}$ estimates of screened SPARK phenotypes ($N_{pheno}$=47).** GCTA $h^2_{SNP}$ estimates for **(A)** categorical phenotypes are shown for untransformed scores (light green) and deviance residuals (dark green). GCTA $h^2_{SNP}$ estimates for **(B)** continuous phenotypes are shown for untransformed scores (light green) and rank-transformed residuals (dark green). The error bars represent standard errors. GCTA $h^2_{SNP}$ estimates were adjusted for sex, age, age squared, and ten ancestry-informative principal components. For a detailed description of SPARK phenotypes see Supplementary Table 1.

Abbreviations: ADHD (attention deficit hyperactivity disorder), DCDQ (Developmental Coordination Disorder Questionnaire), $h^2_{SNP}$ (single nucleotide polymorphism-based heritability), OCD (obsessive-compulsive disorder), ODD (oppositional defiant disorder), PCD (pragmatic communication disorder), RBSR (Repetitive Behaviour Scale-Revised), SCQ (Social Communication Questionnaire).

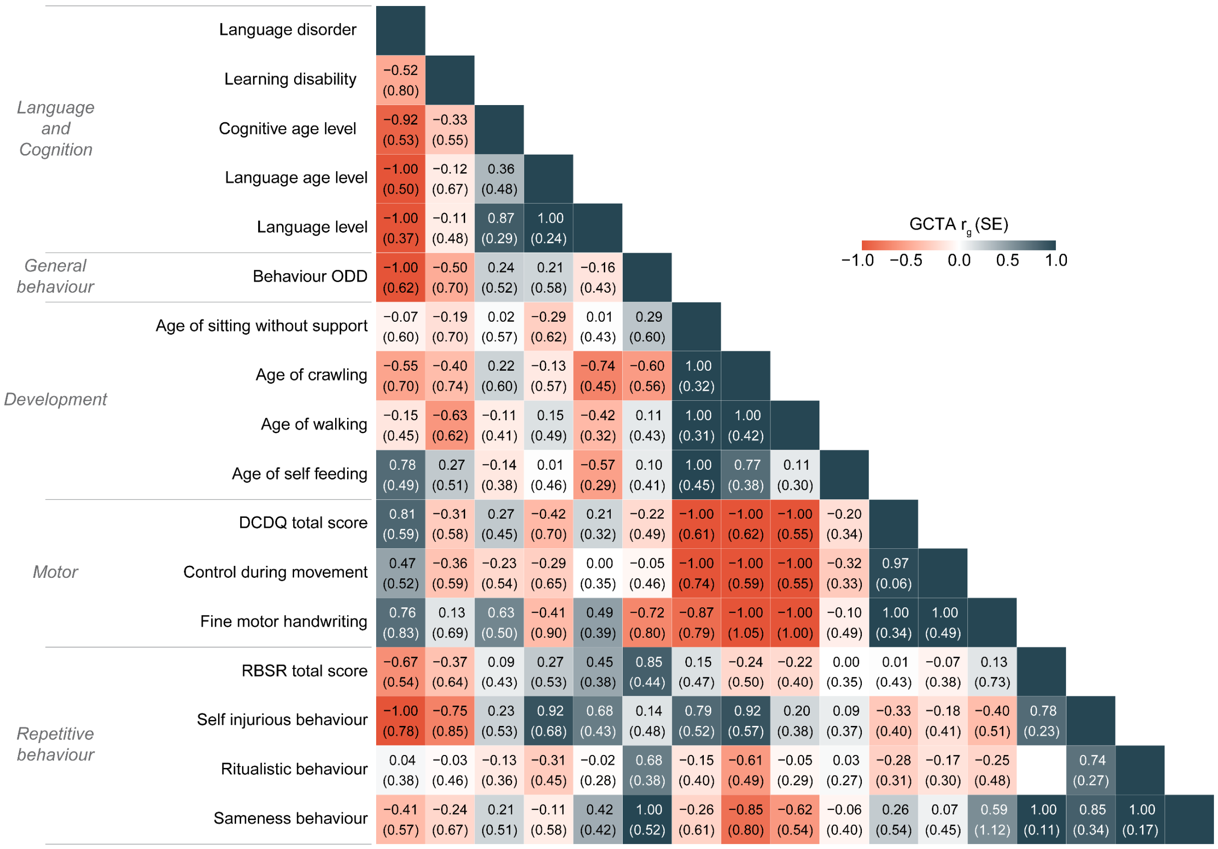

**Supplementary Figure 4. Genetic correlations ($r_g$) across phenotypes in SPARK.** Estimated GCTA genetic correlations based on transformed scores (deviance residuals for categorical phenotypes and rank-transformed residuals for continuous, $N_{pheno}$=17 with GCTA $h^2_{SNP}$ $p \leq 0.1$), adjusted for sex, age, age squared, and ten ancestry-informative principal components. Abbreviations: DCDQ (Developmental Coordination Disorder Questionnaire), ODD (oppositional defiant disorder), RBSR (Repetitive Behaviour Scale-Revised).

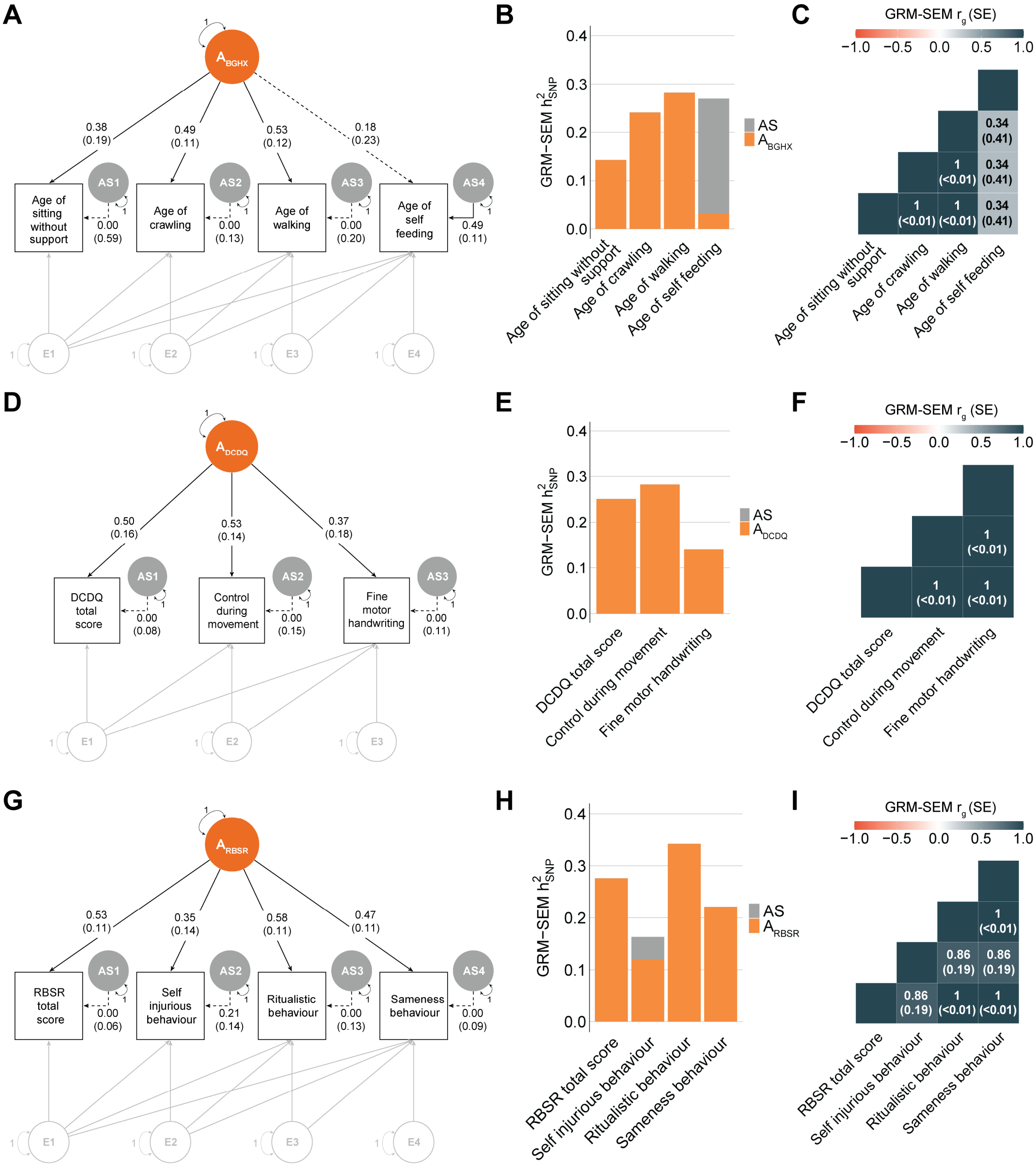

**Supplementary Figure 5. Proxy measure identification in SPARK.** Hybrid GRM-SEM one-factor independent pathway/Cholesky (IPC) models were fitted across scores from the same questionnaire to identify shared genetic influences. **(A)** Path diagram, **(B)** standardised genetic variance plot and **(C)** genetic correlations for scores of the Background History Child Questionnaire (BGHX). **(D)** Path diagram, **(E)** standardised genetic variance plot and **(F)** genetic correlations for items across the Developmental Coordination Disorder Questionnaire (DCDQ). **(G)** Path diagram, **(H)** standardised genetic variance plot and **(I)** genetic correlations for items across the Repetitive Behaviour Scale-Revised (RBSR). **(A,D,G)** Observed measures are represented by squares and latent factors by circles. Single-headed arrows (paths) define relationships between variables. Dotted and solid paths represent factor loadings with $p$>0.05 and $p$≤0.05 respectively. The genetic part of the model has been modelled using an Independent Pathway model, and the residual part using a Cholesky model (grey). **(C,F,I)** SEs for GRM-SEM $h^2_{SNP}$ contributions have been omitted for clarity.

*Abbreviations:* $h^2_{SNP}$ (single nucleotide polymorphism-based heritability), $r_g$ (genetic correlation).

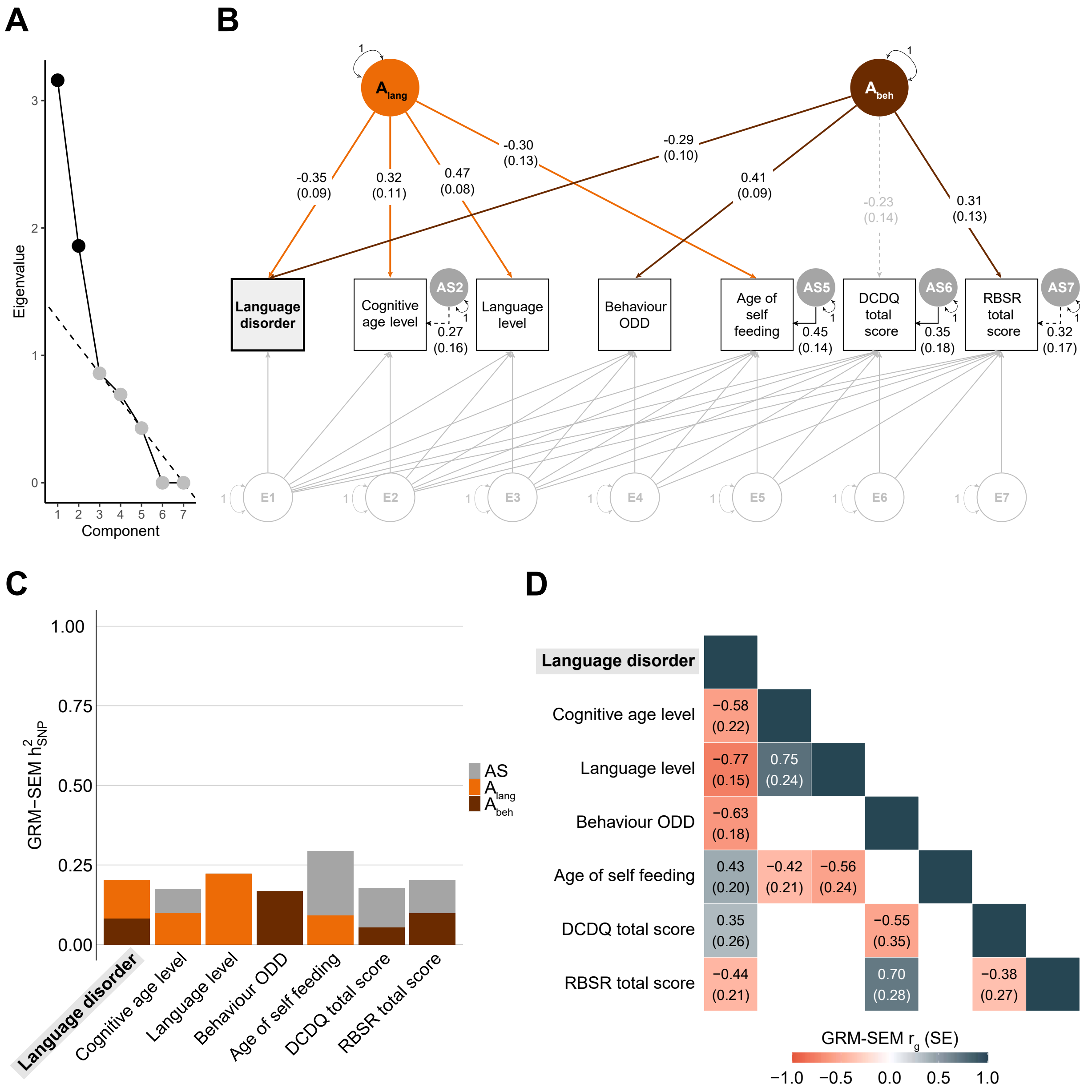

**Supplementary Figure 6. Best-fitting two-factor GRM-SEM model for the language disorder ($S_{DLD}$) set in SPARK.(A)** Scree plot based on the eigenvalue decomposition of genetic correlations derived from a GRM-SEM Cholesky model, depicting the number of expected shared genetic factors (in black) according to an optimal coordinate criterion. The dashed line predicts the "scree" of the plot (grey). **(B)** Path diagram of the best-fitting model (hybrid IPC model with two independent genetic factors: $A_{lang}$, $A_{beh}$) describing genomic covariance in phenotypes linked to language disorder (subset $S_{DLD}$). Observed measures are represented by squares and latent factors by circles. Single-headed arrows (paths) define relationships between variables. Dotted and solid paths represent factor loadings with p>0.05 and p≤0.05 respectively. The genetic part of the model has been modelled using an Independent Pathway model, and the residual part using a Cholesky model (grey). **(C)** Corresponding standardised genetic variance (GRM-SEM $h^2_{SNP}$) plot. SEs for GRM-SEM $h^2_{SNP}$ contributions have been omitted for clarity. **(D)** Corresponding correlogram of genetic correlations. Abbreviations: DCDQ (Developmental Coordination Disorder Questionnaire), $h^2_{SNP}$ (single nucleotide polymorphism-based heritability), IPC (Independent Pathway-Cholesky GRM-SEM model), ODD (Oppositional Defiant Disorder), RBSR (Repetitive Behaviours Scale-Revised), $r_g$ (genetic correlation).

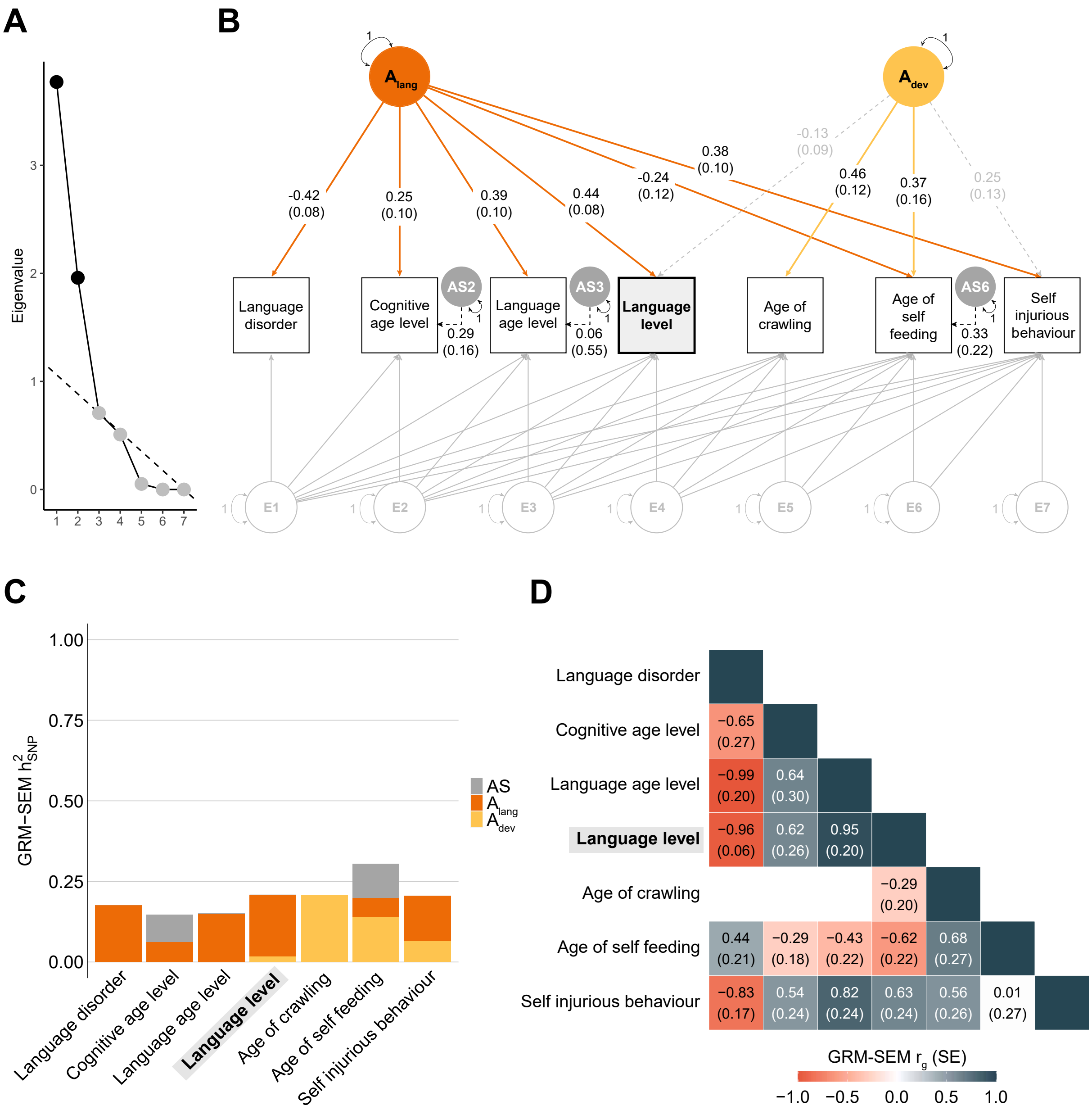

**Supplementary Figure 7. Best-fitting two-factor GRM-SEM model for the language level ($S_{LL}$) set in SPARK. (A)** Scree plot based on the eigenvalue decomposition of genetic correlations derived from a GRM-SEM Cholesky model, depicting the number of expected shared genetic factors (in black) according to an optimal coordinate criterion. The dashed line predicts the “scree” of the plot (grey). **(B)** Path diagram of the best-fitting model (hybrid IPC model with two independent genetic factors: $A_{lang}$, $A_{dev}$) describing genomic covariance in phenotypes genetically correlated to language level (subset $S_{LL}$). Observed measures are represented by squares and latent factors by circles. Single-headed arrows (paths) define relationships between variables. Dotted and solid paths represent factor loadings with $p>0.05$ and $p\leq0.05$ respectively. The genetic part of the model has been modelled using an Independent Pathway model, and the residual part using a Cholesky model (grey). **(C)** Corresponding standardised genetic variance (GRM-SEM $h^2_{SNP}$) plot. SEs for GRM-SEM $h^2_{SNP}$ contributions have been omitted for clarity. **(D)** Corresponding correlogram of genetic correlations.
Abbreviations: $h^2_{SNP}$ (single nucleotide polymorphism-based heritability), IPC (Independent Pathway-Cholesky GRM-SEM model), $r_g$ (genetic correlation).

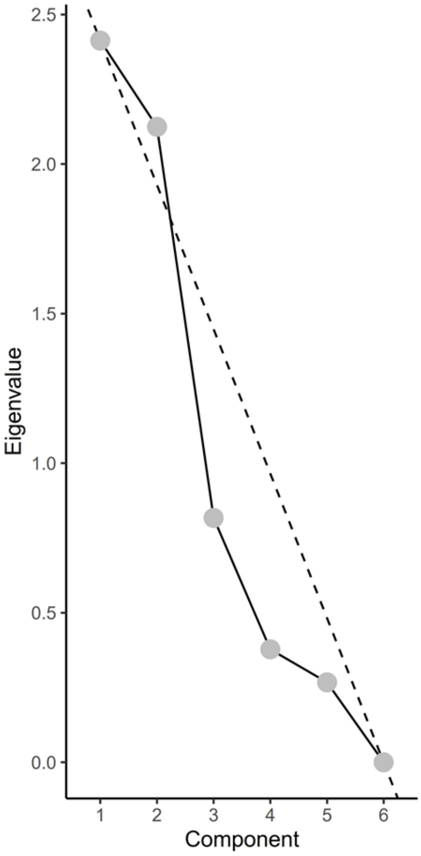

**Supplementary Figure 8. Scree plot for the age of crawling ($S_{CRL}$) set in SPARK.** Scree plot based on the eigenvalue decomposition of genetic correlations derived from a GRM-SEM Cholesky model, depicting the number of expected shared genetic factors (in black) according to an optimal coordinate criterion. The dashed line predicts the "scree" of the plot (grey). For this phenotype subset ($S_{CRL}$), the number of genetic factors could not be predicted by the optimal coordinate criterion.

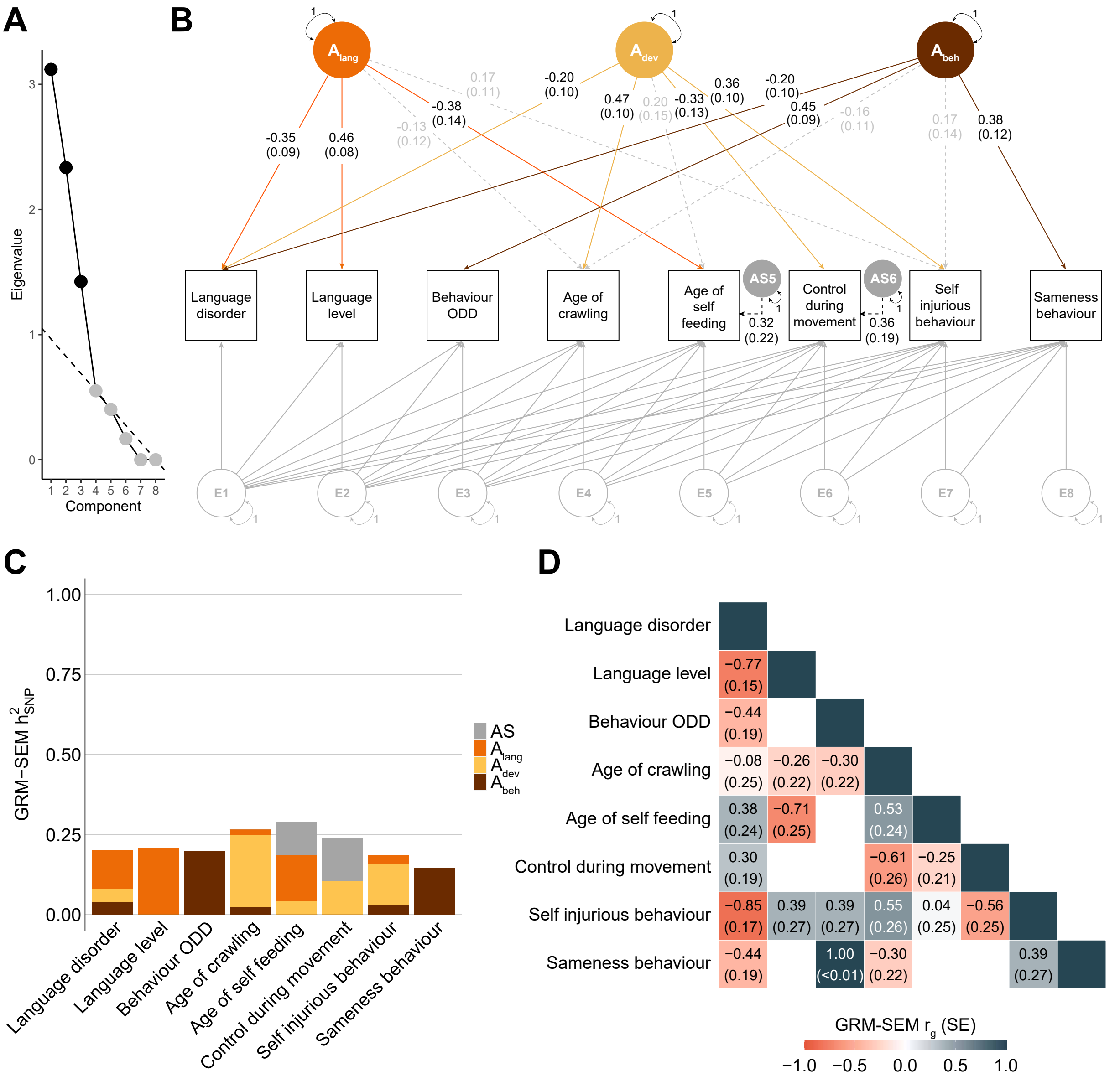

**Supplementary Figure 9. Best-fitting three-factor GRM-SEM model for the combined ($S_{ALL}$) (A)** Scree plot based on the eigenvalue decomposition of genetically derived correlation matrix from a GRM-SEM Cholesky model, depicting the number of expected shared genetic factors (in black) according to an optimal coordinate criterion. The dashed line predicts the "scree" of the plot (grey). **(B)** Path diagram of the best-fitting model (hybrid IPC model with three independent genetic factors: $A_{lang}$, $A_{dev}$, $A_{beh}$) describing genomic covariance in the combined $S_{ALL}$ set ($S_{DLD}$, $S_{LL}$, $S_{CRL}$). Observed measures are represented by squares and latent variables by circles. Dotted and solid single-headed arrows (factor loadings) define relationships between variables with $p$>0.05 and $p$≤0.05, respectively. The genetic part of the model has been modelled using an Independent Pathway model, and the residual part using a Cholesky model (grey). **(C)** Corresponding standardised genetic variance (GRM-SEM $h^2_{SNP}$) plot. SEs for GRM-SEM $h^2_{SNP}$ contributions have been omitted for clarity. **(D)** Corresponding correlogram of genetic correlations.
*Abbreviations:* $h^2_{SNP}$ (single nucleotide polymorphism-based heritability), IPC (Independent Pathway-Cholesky GRM-SEM model), ODD (Oppositional Defiant Disorder), $r_g$ (genetic correlation).

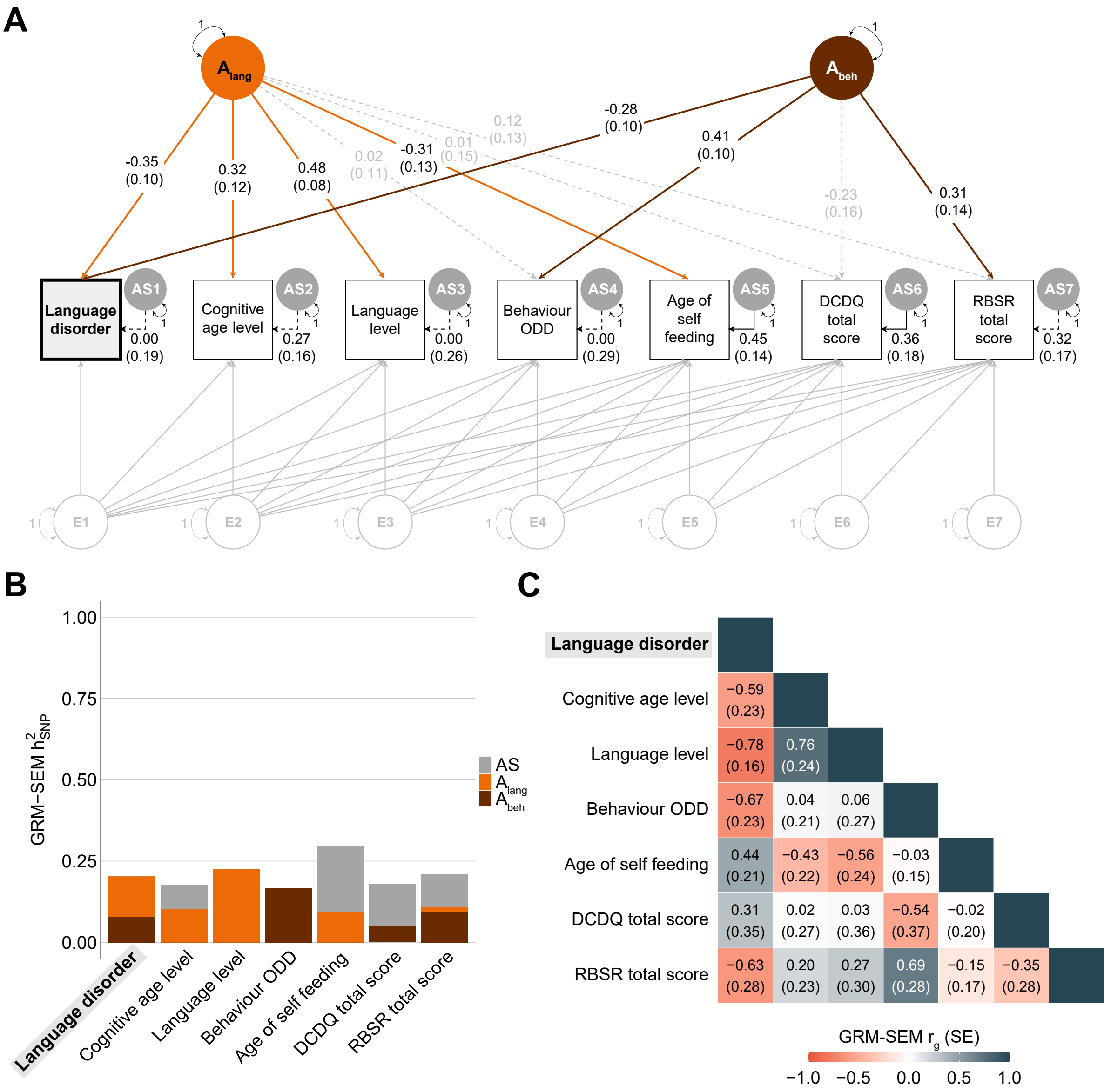

**Supplementary Figure 10. Bi-factor GRM-SEM model for the language disorder ($S_{DLD}$) set in SPARK. (A)** Path diagram of a hybrid IPC bi-factor model with two independent genetic factors ($A_{lang}$, $A_{beh}$) describing genomic covariance in phenotypes linked to language disorder (subset $S_{DLD}$). Observed measures are represented by squares and latent factors by circles. Single-headed arrows (paths) define relationships between variables. Dotted and solid paths represent factor loadings with *p*>0.05 and *p*≤0.05 respectively. The genetic part of the model has been modelled using an Independent Pathway model, and the residual part using a Cholesky model (grey). **(C)** Corresponding standardised genetic variance (GRM-SEM $h^2_{SNP}$) plot. SEs for GRM-SEM $h^2_{SNP}$ contributions have been omitted for clarity. **(D)** Corresponding correlogram of genetic correlations.
*Abbreviations:* DCDQ (Developmental Coordination Disorder Questionnaire), $h^2_{SNP}$ (single nucleotide polymorphism-based heritability), IPC (Independent Pathway-Cholesky GRM-SEM model), ODD (Oppositional Defiant Disorder), RBSR (Repetitive Behaviours Scale-Revised), $r_g$ (genetic correlation).

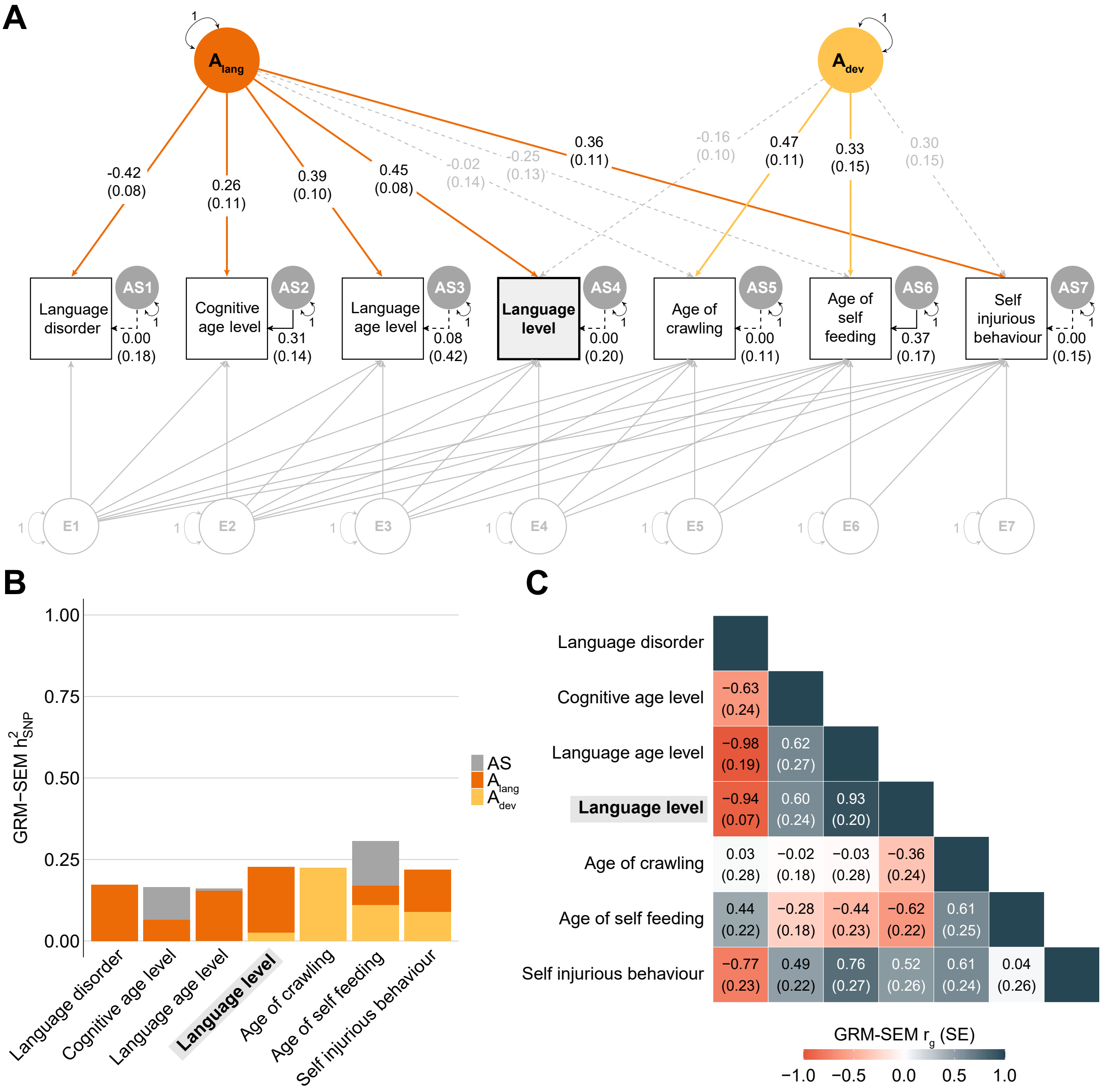

**Supplementary Figure 11. Bi-factor GRM-SEM model for the language level ($S_{LL}$) set in SPARK. (A)** Path diagram of a hybrid IPC bi-factor model with two independent genetic factors ($A_{lang}$, $A_{dev}$) describing genomic covariance in phenotypes genetically correlated to language level (subset $S_{LL}$). Observed measures are represented by squares and latent factors by circles. Single-headed arrows (paths) define relationships between variables. Dotted and solid paths represent factor loadings with $p$>0.05 and $p$≤0.05 respectively. The genetic part of the model has been modelled using an Independent Pathway model, and the residual part using a Cholesky model (grey). **(C)** Corresponding standardised genetic variance (GRM-SEM $h^2_{SNP}$) plot. SEs for GRM-SEM $h^2_{SNP}$ contributions have been omitted for clarity. **(D)** Corresponding correlogram of genetic correlations.

*Abbreviations:* $h^2_{SNP}$ (single nucleotide polymorphism-based heritability), IPC (Independent Pathway-Cholesky GRM-SEM model), $r_g$ (genetic correlation).

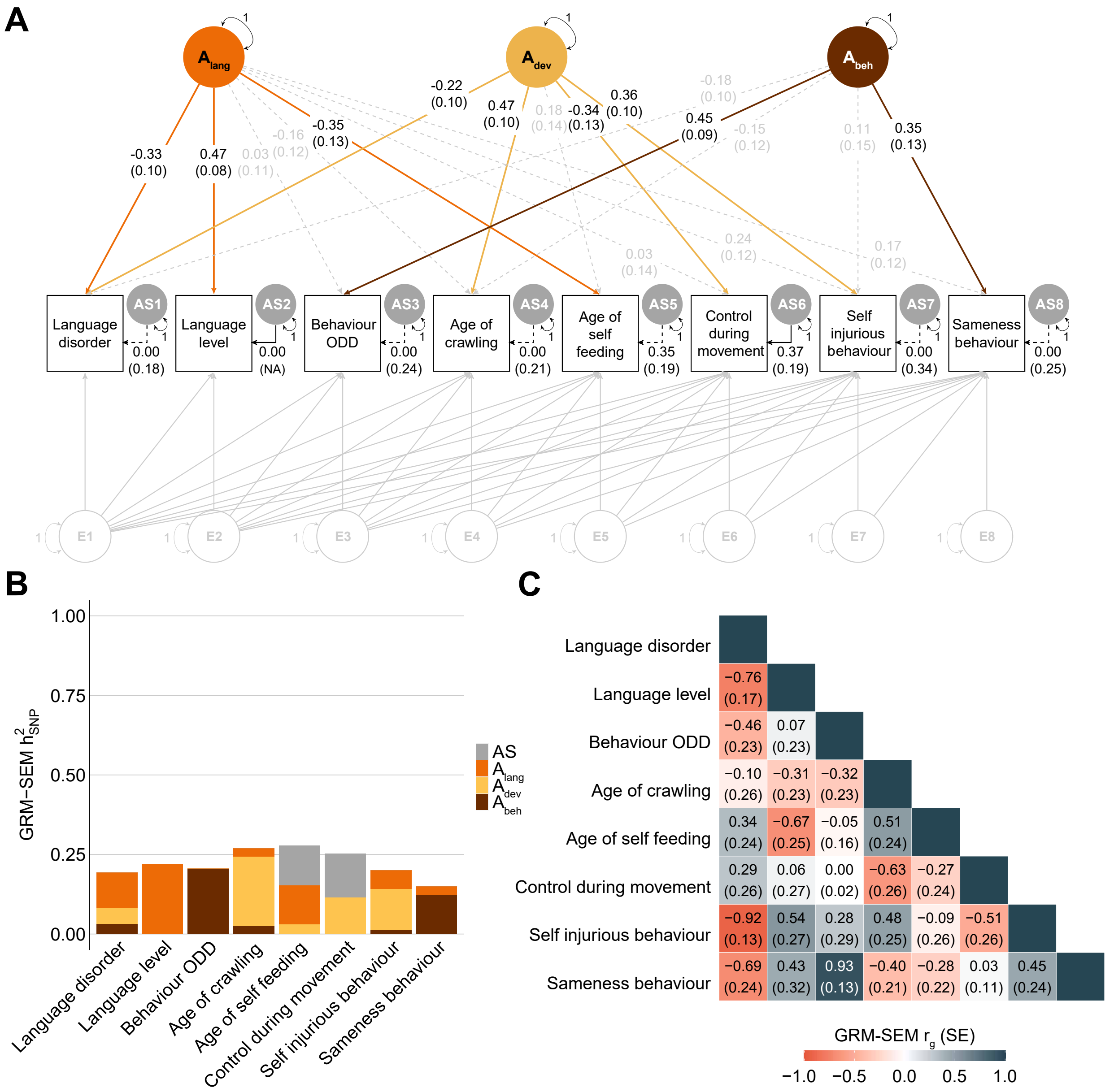

**Supplementary Figure 12. Bi-factor GRM-SEM model for the combined ($S_{ALL}$) set in SPARK.(A)** Path diagram of a hybrid IPC bi-factor model with three independent genetic factors ($A_{lang}$, $A_{dev}$, $A_{beh}$) describing genomic covariance in the combined $S_{ALL}$ data set ($S_{DLD}$, $S_{LL}$, $S_{CRL}$). Observed measures are represented by squares and latent factors by circles. Single-headed arrows (paths) define relationships between variables. Dotted and solid paths represent factor loadings with $p$>0.05 and $p$≤0.05 respectively. The genetic part of the model has been modelled using an Independent Pathway model, and the residual part using a Cholesky model (grey). **(C)** Corresponding standardised genetic variance (GRM-SEM $h^2_{SNP}$) plot. SEs for GRM-SEM $h^2_{SNP}$ contributions have been omitted for clarity. **(D)** Corresponding correlogram of genetic correlations.

*Abbreviations:* $h^2_{SNP}$ (single nucleotide polymorphism-based heritability), IPC (Independent Pathway-Cholesky GRM-SEM model), ODD (Oppositional Defiant Disorder), $r_g$ (genetic correlation).

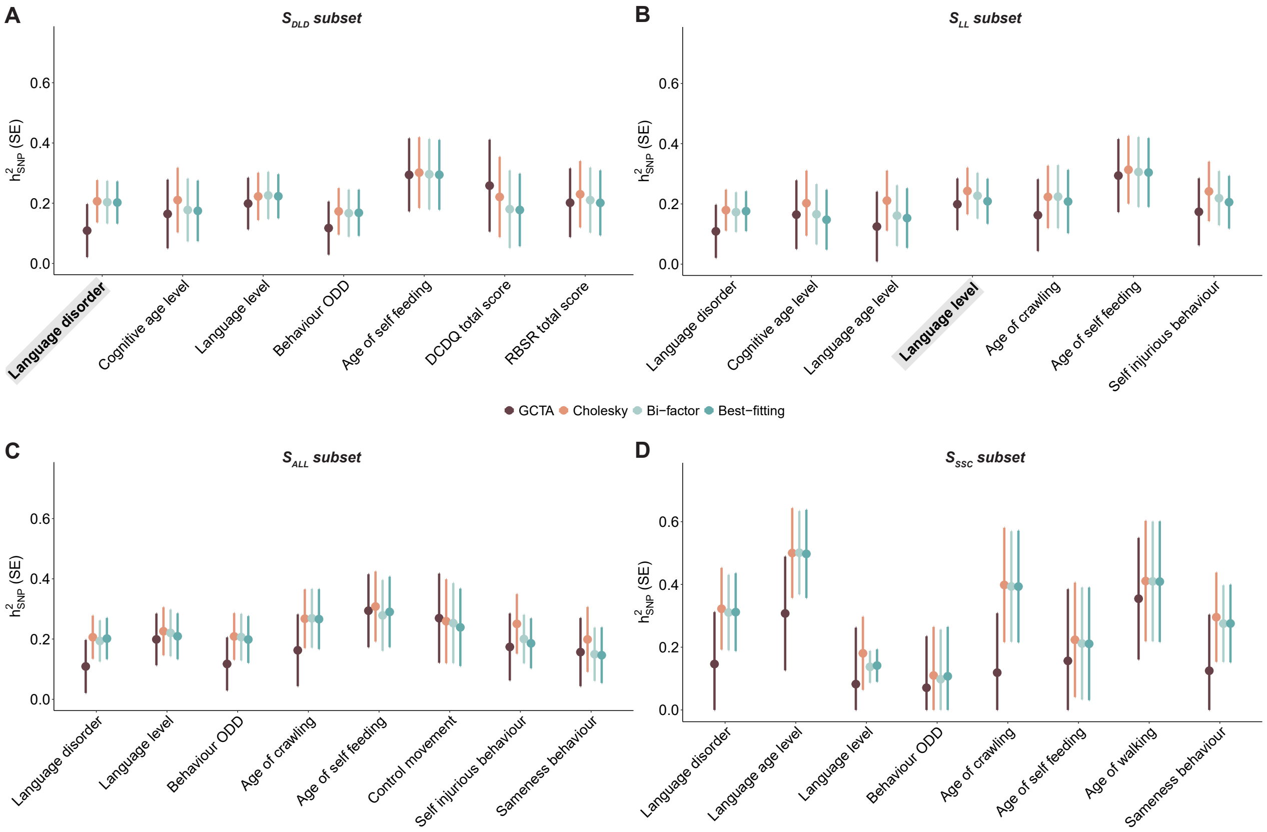

**Supplementary Figure 13. Heritability comparison between GCTA and GRM-SEM.** $h^2_{SNP}$ comparison across structural models fitted to the **(A)** language disorder ($S_{DLD}$), **(B)** language level ($S_{LL}$) and **(C)** combined set ($S_{ALL}$) in SPARK and **(D)** the follow-up set in the SSC ($S_{SSC}$), based on univariate GCTA as well as GRM-SEM Cholesky, Bi-factor and best-fitting multifactorial IPC models. Error bars represent standard errors. All analyses are based on transformed scores (categorical phenotypes: deviance residuals; continuous scores: rank-transformed residuals).

*Abbreviations:* DCDQ (Developmental Coordination Disorder Questionnaire), $h^2_{SNP}$ (single nucleotide polymorphism-based heritability), IPC (Independent Pathway-Cholesky GRM-SEM model), ODD (Oppositional Defiant Disorder), RBSR (Repetitive Behaviours Scale-Revised).

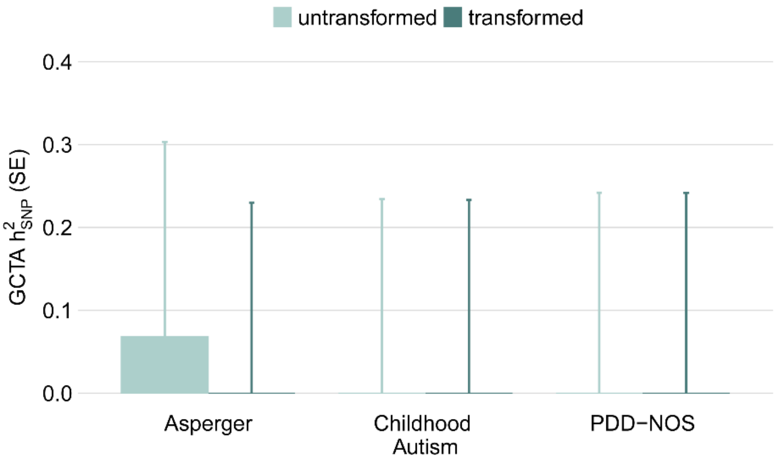

**Supplementary Figure 14. GCTA $h^2_{SNP}$ estimates of dichotomised ASD subcategories in SPARK.** The error bars represent standard errors. GCTA $h^2_{SNP}$ estimates for transformed scores were adjusted for sex, age, age squared, and ten ancestry-informative principal components. From a total sample of 5,331 ASD unrelated probands with phenotypic and genotyping information, data from 716 probands with Asperger diagnosis were available (566 males, 150 females), for Childhood Autism data for 624 probands were available (509 males, 115 females) and for PDD-NOS data for 414 probands were available (334 males, 80 females). Contrasts were coded as follows: Asperger (Asperger=1, Childhood Autism=0, PDD-NOS=0, not-available=NA), Childhood Autism (Asperger=0, Childhood Autism=1, PDD-NOS=0, not-available=NA) and PDD-NOS (Asperger=0, Childhood Autism=0, PDD-NOS=1, not-available=NA). *Abbreviations:* $h^2_{SNP}$ (single nucleotide polymorphism-based heritability), PDD-NOS (Pervasive Developmental Disorder Not Otherwise Specified).

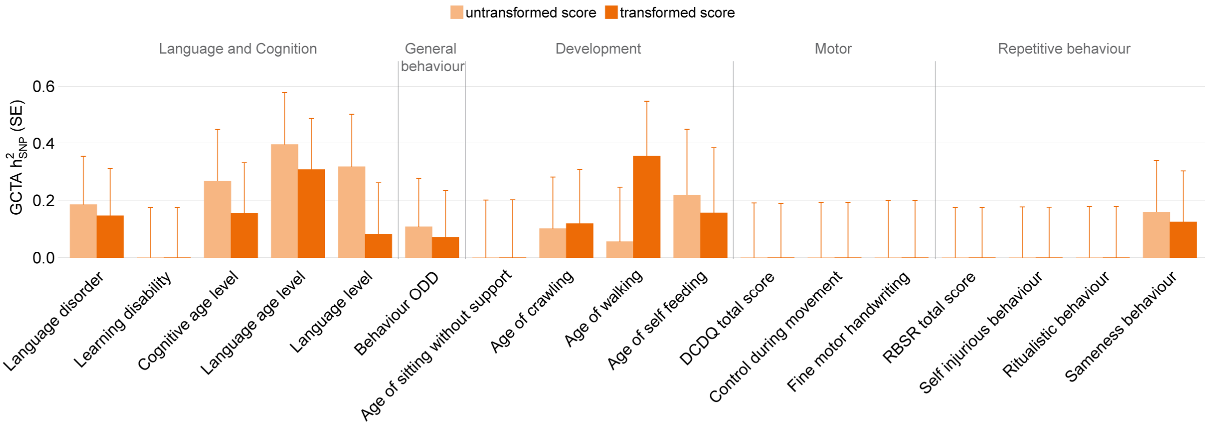

**Supplementary Figure 15. GCTA $h^2_{SNP}$ estimates of Simons Simplex Collection (SSC) phenotypes (follow-up).** The error bars represent standard errors. GCTA $h^2_{SNP}$ estimates for categorical phenotypes are shown for untransformed scores (light orange) and deviance residuals (dark orange). GCTA $h^2_{SNP}$ estimates for continuous phenotypes are shown for untransformed scores (light orange) and rank-transformed residuals (dark orange). GCTA $h^2_{SNP}$ estimates were adjusted for sex, age, age squared, and ten ancestry-informative principal components. For a detailed description of SSC phenotypes see Supplementary Table 2. Abbreviations: DCDQ (Developmental Coordination Disorder Questionnaire), $h^2_{SNP}$ (single nucleotide polymorphism-based heritability), ODD (oppositional defiant disorder), RBSR (Repetitive Behaviour Scale-Revised).

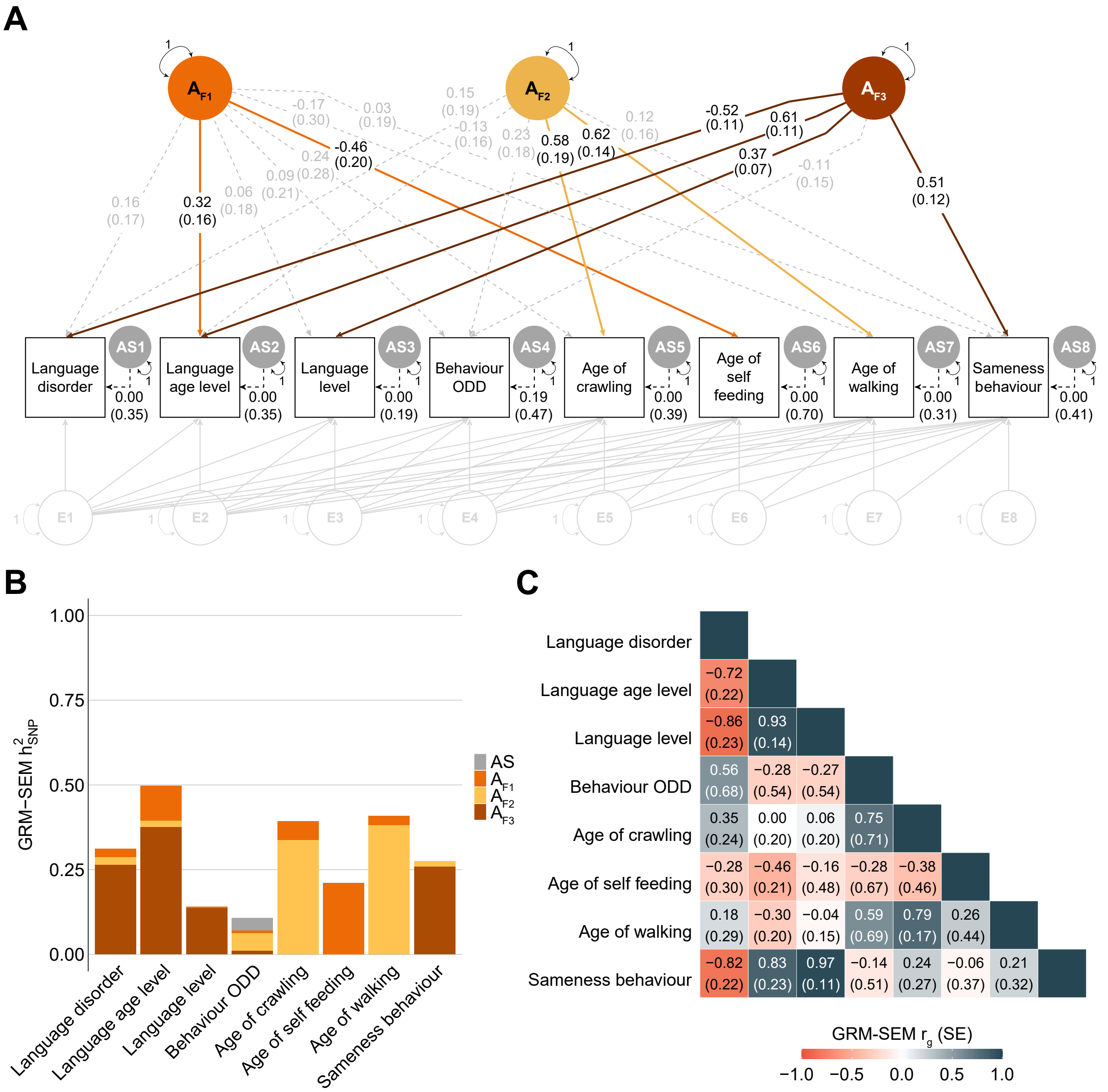

**Supplementary Figure 16. Follow-up bi-factor GRM-SEM model in the SSC ($S_{SSC}$).(A)** Path diagram of a hybrid IPC bi-factor model with three independent genetic factors describing genomic covariance in largely comparable phenotypes as studied in the combined $S_{ALL}$ SPARK set. Observed measures are represented by squares and latent variables by circles. Dotted and solid single-headed arrows (factor loadings) define relationships between variables with $p$>0.05 and $p$≤0.05, respectively. The genetic part of the model has been modelled using an Independent Pathway model, and the residual part using a Cholesky model (grey). **(B)** Corresponding standardised genetic variance (GRM-SEM $h^2_{SNP}$) plot. SEs for GRM-SEM $h^2_{SNP}$ contributions have been omitted for clarity. **(C)** Corresponding correlogram of genetic correlations.

*Abbreviations:* $h^2_{SNP}$ (Single nucleotide polymorphism-based heritability), IPC (Independent Pathway-Cholesky GRM-SEM model), $r_g$ (genetic correlation).

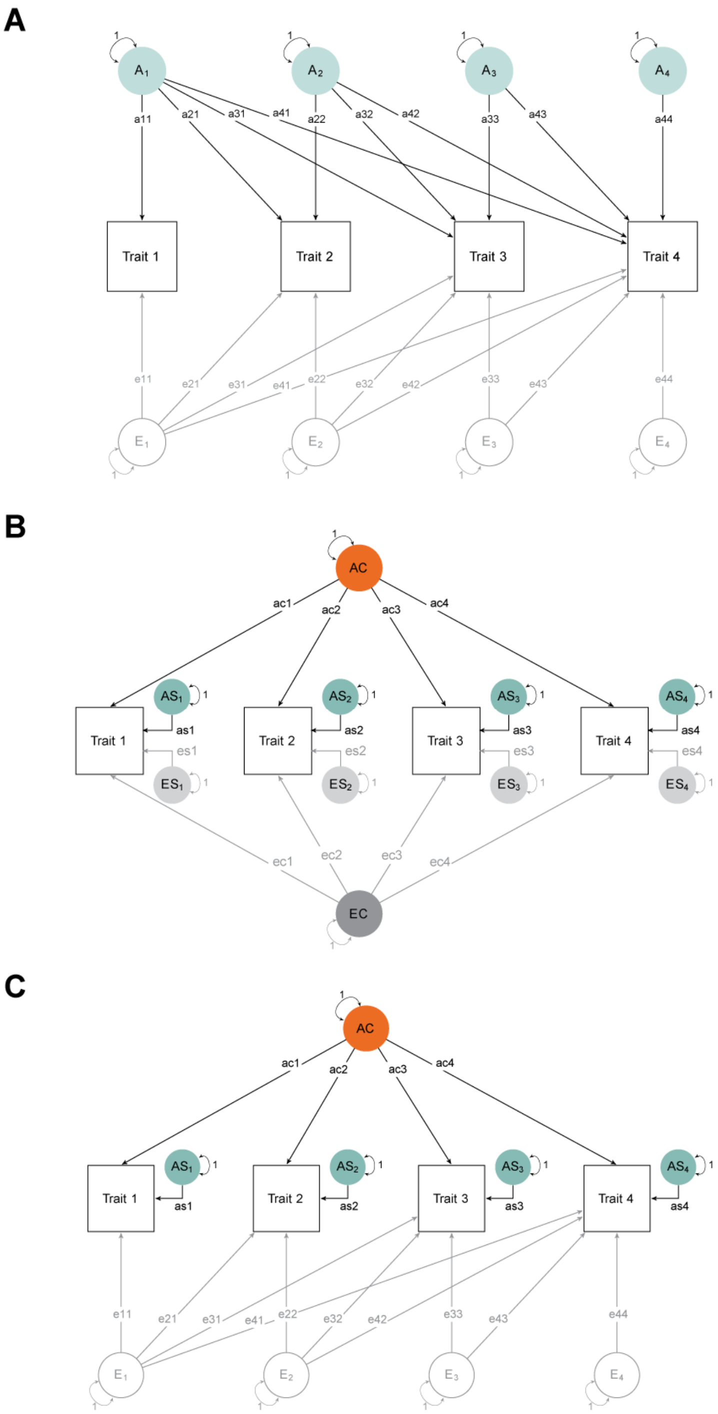

**Supplementary Figure 17. GRM-SEM multivariate model types. (A)** Cholesky decomposition model (saturated) **(B)** Independent Pathway (IP) model. **(C)** Hybrid Independent Pathway-Cholesky (IPC) model, where the genetic variance is modelled with an IP structure and the residual variance with a Cholesky decomposition.

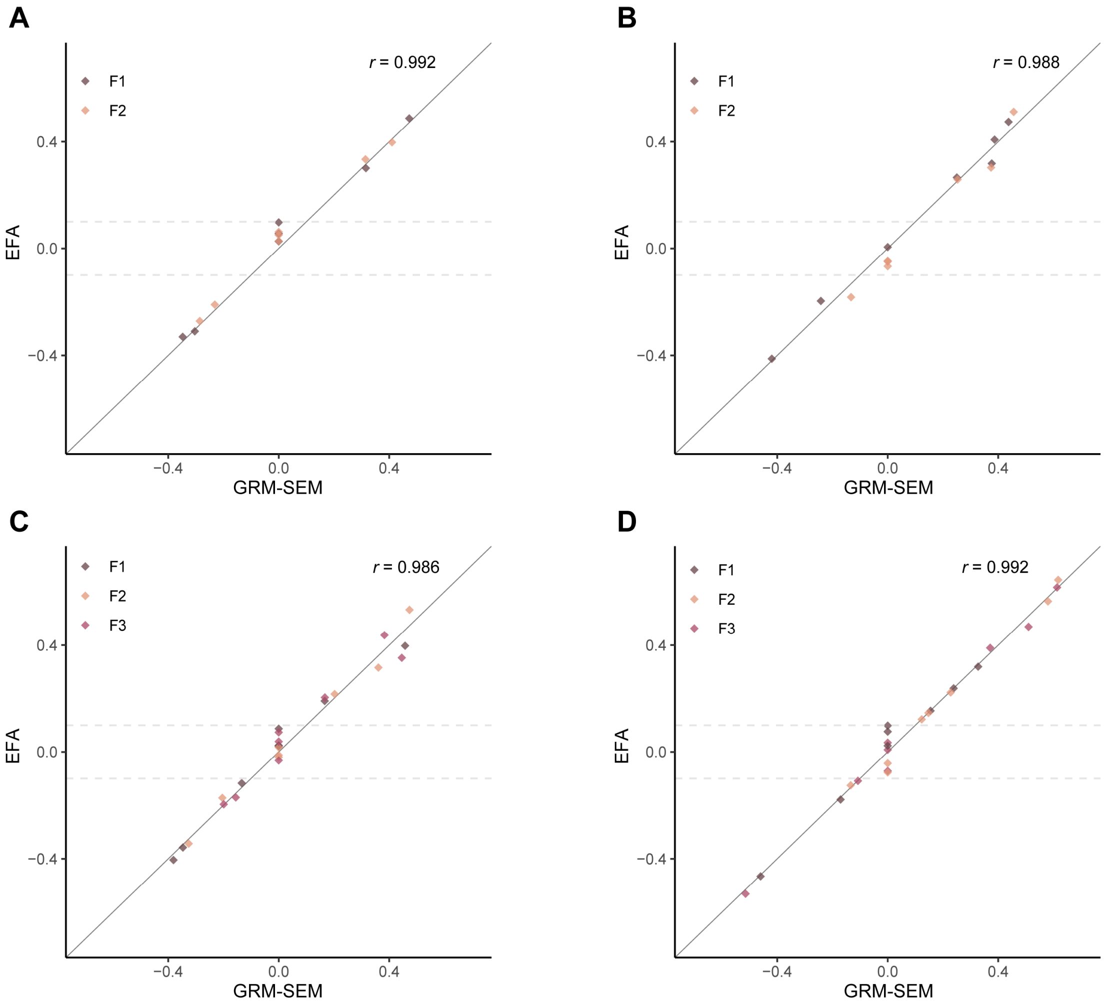

**Supplementary Figure 18. Comparison of factor loadings estimated with EFA *lavaan* and GRM-SEM.** Factor loadings for genetic factors (F1-3) were compared between exploratory factor analysis (EFA) *lavaan* software (varimax rotation, DWLS algorithm) and GRM-SEM models fitted to the **(A)** language disorder subset ($S_{DLD}$), **(B)** language level subset ($S_{LL}$) and **(C)** combined set ($S_{ALL}$) in SPARK as well as to the **(D)** follow-up set in the SSC ($S_{SSC}$). The correlation coefficient (*r*, Pearson) between EFA and GRM-SEM estimates is shown in the graph for each subset.

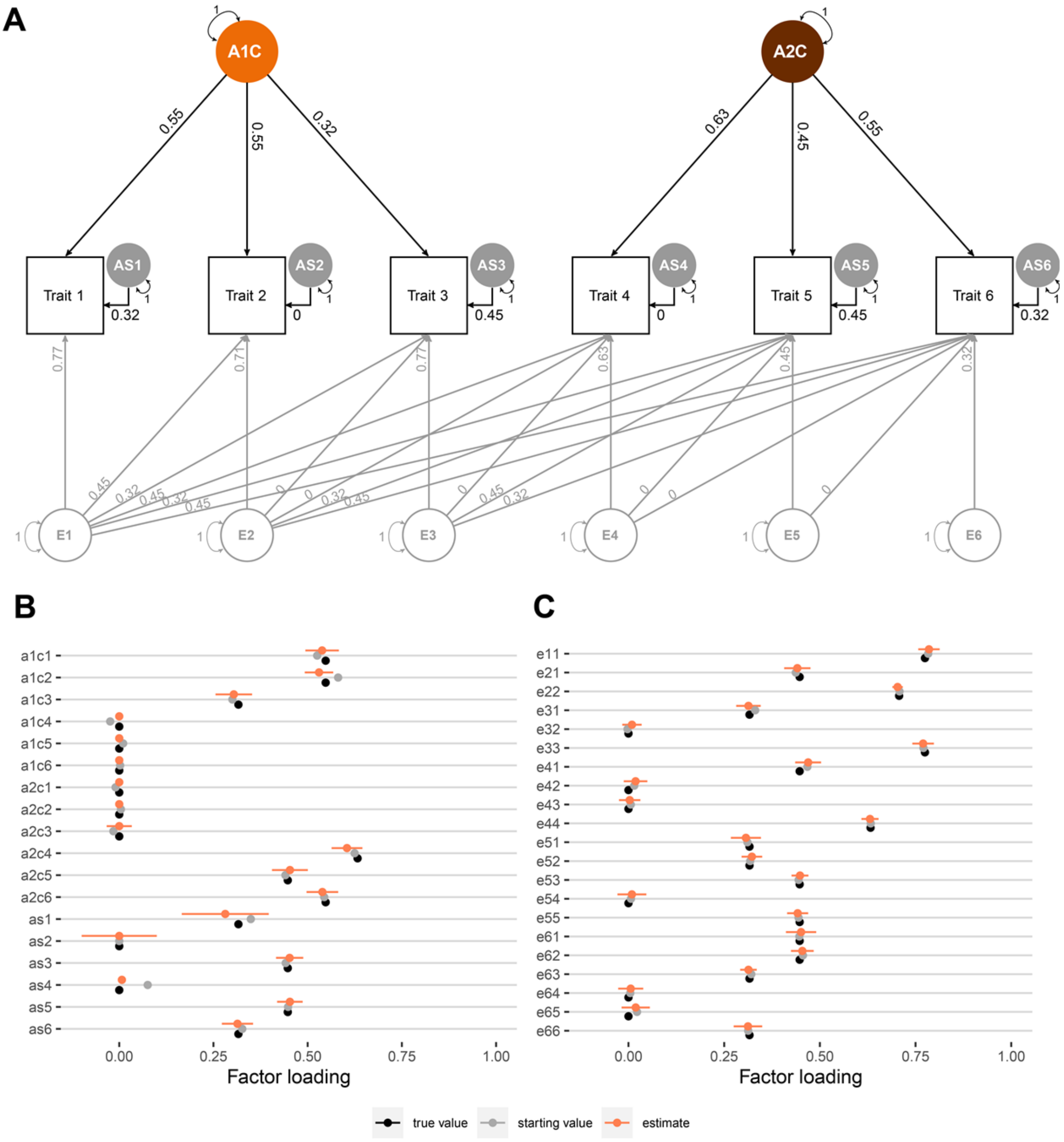

**Supplementary Figure 19. GRM-SEM simulations of a six-variate trait with two independent common genetic factors without cross-loadings. (A)** Path diagram of the simulated six-variate trait, assuming two independent genetic factors (A1C and A2C) without cross-loading, based on 2,000 individuals per trait and (for simplicity) 5,000 causal loci. **(B)** Genetic and **(C)** residual factor loadings with true values, median starting values and median estimated values (± Empirical SE) across 20 simulations based on a data-driven genomic covariance modelling approach (Figure 1B). The number of genetic factors was based on the eigenvalue decomposition of genetic correlations derived from a GRM-SEM Cholesky model. IPC starting values and constraints for the genetic part were obtained from an Exploratory Factor Analysis (EFA) *lavaan* model fitted to the Cholesky-estimated genetic variance/covariance matrix. EFA-predicted genetic factor loadings |λ|< 0.1 were constrained to zero. Starting values for the residual part were informed by the pre-fitted Cholesky model.

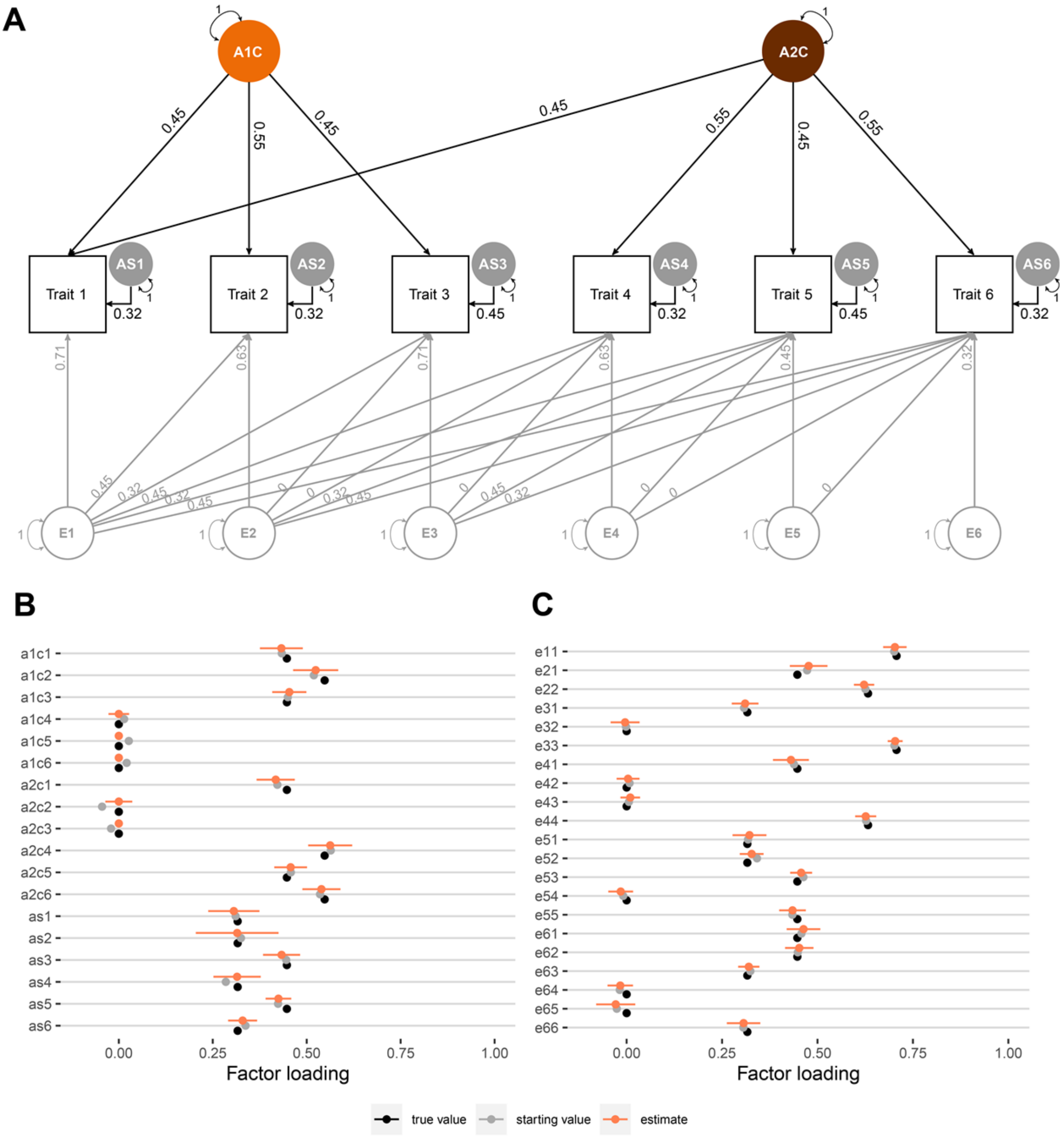

**Supplementary Figure 20. GRM-SEM simulations of a six-variate trait with two independent genetic factors with cross-loading. (A)** Path diagram of a six-variate trait, assuming two genetic factors (A1C and A2C) with cross-loading, based on 2,000 individuals per trait and (for simplicity) 5,000 causal loci. **(B)** Genetic and **(C)** residual factor loadings with true values, median starting values and median estimated values (± Empirical SE) across 20 simulations based on a data-driven genomic covariance modelling approach (Figure 1B). The number of genetic factors was based on the eigenvalue decomposition of genetic correlations derived from a GRM-SEM Cholesky model. IPC starting values and constraints for the genetic part were obtained from an Exploratory Factor Analysis (EFA) *lavaan* model fitted to the Cholesky-estimated genetic variance/covariance matrix. EFA-predicted genetic factor loadings |λ|<0.1 were constrained to zero. Starting values for the residual part were informed by the pre-fitted Cholesky model.

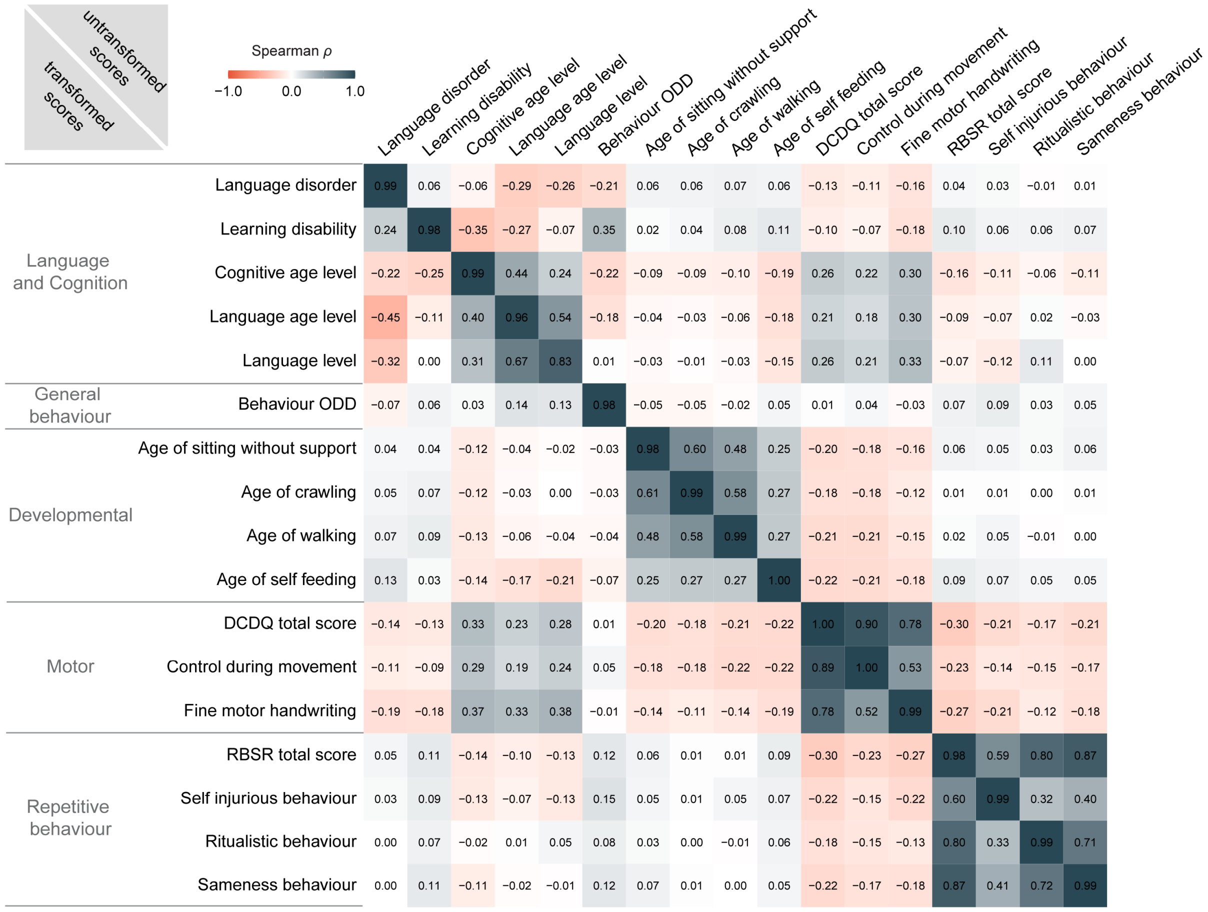

**Supplementary Figure 21. Phenotypic correlations of ASD phenotypes in SPARK.** Figure shows phenotypic correlations of ASD SPARK phenotypes for which the $h^2_{SNP}$ met a significance threshold of $p \leq 0.1$, including transformed scores (lower triangle) and untransformed scores (upper triangle). The diagonal shows the correlation between the untransformed and transformed scores. Transformed categorical phenotypes are based on deviance residuals, transformed continuous phenotypes on rank-transformed residuals. Transformed scores were adjusted for sex, age, age squared, and ten ancestry-informative principal components.

*Abbreviations:* DCDQ (Developmental Coordination Disorder Questionnaire), $h^2_{SNP}$ (single nucleotide polymorphism-based heritability), ODD (oppositional defiant disorder), RBSR (Repetitive Behaviour Scale-Revised).
